# Clinical and immunological features of SARS-CoV-2 breakthrough infections in vaccinated individuals requiring hospitalization

**DOI:** 10.1101/2022.02.14.22270857

**Authors:** Giulia Lamacchia, Alessio Mazzoni, Michele Spinicci, Anna Vanni, Lorenzo Salvati, Benedetta Peruzzi, Sara Bencini, Manuela Capone, Alberto Carnasciali, Parham Farahvachi, Arianna Rocca, Seble Tekle Kiros, Lucia Graziani, Lorenzo Zammarchi, Jessica Mencarini, Maria Grazia Colao, Roberto Caporale, Francesco Liotta, Lorenzo Cosmi, Gian Maria Rossolini, Alessandro Bartoloni, Laura Maggi, Francesco Annunziato

**Affiliations:** Department of Experimental and Clinical Medicine, University of Florence, Florence, Italy; Infectious and Tropical Diseases Unit, Careggi University Hospital, Florence, Italy; Flow cytometry diagnostic center and immunotherapy, Careggi University Hospital, Florence, Italy; Microbiology and Virology Unit, Careggi University Hospital, Florence, Italy; Immunology and Cell Therapy Unit, Careggi University Hospital, Florence, Italy

## Abstract

**Background:** Waning immunity and the surge of SARS-CoV-2 variants are responsible for breakthrough infections, i.e. infections in fully vaccinated individuals. Although the majority of vaccinated infected subjects reports mild or no symptoms, some others require hospitalization. The clinical and immunological features of vaccinated hospitalized COVID-19 patients are currently unknown.

**Methods:** 29 unvaccinated and 36 vaccinated hospitalized COVID-19 patients were prospectively enrolled and clinical and laboratory data. Immunophenotyping of leukocytes’ subsets, T and B cell SARS-CoV-2 specific responses were evaluated via flow cytometry. Anti-IFN-α autoantibodies were measured via ELISA.

**Results:** Despite vaccinated patients were older and with more comorbidities, unvaccinated subjects showed higher levels of pro-inflammatory markers, more severe disease and increased mortality rate. Accordingly, they presented significant alterations in the circulating leukocyte composition, typical of severe COVID-19. Vaccinated patients displayed higher levels of anti-Spike IgGs and Spike-specific B cells. Of all participants, survivors showed higher levels of anti-Spike IgGs and S-specific CD4+ T cells than non-survivors. At hospital admission, 6 out of 65 patients (9.2%) displayed high serum concentrations of autoantibodies targeting IFN-α. Remarkably, 3 were unvaccinated and eventually died, while the other 3 were vaccinated and survived.

**Conclusion:** Despite more severe pre-existing clinical conditions, vaccinated patients have good outcome. A rapid activation of anti-SARS-CoV-2-specific immunity is fundamental for the resolution of the infection. Therefore, prior immunization through vaccination provides a significant contribute to prevention of disease worsening and can even overcome the presence of high-risk factors (i.e. older age, comorbidities, anti-IFN-α autoantibodies positivity).

## Introduction

As of February 7, 2022, registered COVID-19 cases have exceeded 380 million cases worldwide and more than 5.7 million fatalities (WHO Dashboard). Vaccines’ release in the last weeks of December 2020 changed the perspective of the ongoing COVID-19 pandemic. As vaccination campaigns proceeds towards the vast immunization in most countries, one of the main open matters regards the maintenance of vaccine efficacy over time. Indeed, the progressive waning of antibody titers and the emergence of new viral variants with increased transmissibility and immune escape capability have led to increased numbers of breakthrough infections, i.e. SARS-CoV-2 infections in fully vaccinated individuals (Juthani et al., 2021, Kuhlmann et al.,2022). Published data have demonstrated progressive loss of vaccine efficacy against infection 6 months following the second dose administration, although maintaining substantially conserved efficacy against severe disease (Naaber et al., 2021 Levin et al., 2021). These observations were obtained concomitantly with the global spread of Delta variant. A third vaccine injection was demonstrated to be safe and effective in restoring vaccine efficacy (Mazzoni et al., in press., 2022, Arbel et al., 2021 Bar-On et al.,2021). However, the recent emergence of Omicron variant with enhanced immune escape potential (Ren et al., 2022), has been associated to increased numbers of re-infections and breakthrough infections even after booster administration (Kuhlmann et al., 2022).

Nonetheless, among the many subjects fully vaccinated who test positive for SARS-CoV-2, the striking majority reports mild or no symptoms (Juthani et al., 2021). Indeed, if the reduced antibody neutralization capability predisposes to infection, T cell immunity is substantially conserved against viral variants, thus conferring protection from severe disease (Mazzoni et al., 2022, Tarke et al., 2022). However, in some cases hospitalization is needed, but the predisposing factors for increased risk of severe COVID-19 in vaccinated subjects are currently unknown. Indeed, head-to-head clinical and laboratory in-depth characterizations of vaccinated and unvaccinated hospitalized COVID-19 patients are still missing. This type of analysis may better elucidate the role of vaccination in modifying the disease course. This is of importance, as it could inform about the susceptibility of fully vaccinated subjects to SARS-CoV-2 infection or re-infection, and might also direct attention to the need for updated vaccines, highly specific for the new emerging viral strains.

In the present study, we collected clinical, laboratory and immunological data to characterize vaccinated and unvaccinated subjects infected during the Delta variant wave in Italy, who needed hospital admission. Our results showed that, despite more severe pre-existing clinical conditions, vaccinated subjects presented a more favorable disease course, with higher survival rate than the unvaccinated patients. This clinical observation was supported by immunological findings demonstrating a pattern of significantly altered immune homeostasis in unvaccinated patients. Finally, we found that vaccination provided a rapid activation of anti-SARS-CoV-2 immunity upon infection resulting in a significant positive impact on patients’ survival.

## Results

### Clinical features of vaccinated and unvaccinated COVID-19 patients at hospital admission and throughout hospitalization

A total of 65 patients affected by COVID-19 and admitted to the Careggi University Hospital (Florence, Italy) were enrolled from November to mid-December 2021, a period of time that coincided with the outbreak of B1.617.2 Variant of Concern, commonly known as Delta variant (ECDC Dashboard, weeks from 44-2021 to 49-2021). The cohort was divided into the group of vaccinated subjects (n=36), who received a complete vaccination cycle of any approved viral vector or mRNA SARS-CoV-2 vaccine and possibly a booster dose, and the group of unvaccinated subjects (n=29). All patients tested positive for reverse transcription real-time PCR performed on nasopharyngeal swabs, confirming SARS-CoV-2 infection. Notably, the viral load was comparable between the two cohorts (Supplemental Figure 1). Gender distribution was similar in both groups. As for the vaccinated patients, the mean age was 73 years, significantly higher than unvaccinated individuals (67 years), as shown in Supplemental Table 1. All subjects reported approximately one week between the onset of symptoms and hospital admission, with no differences between the two groups (Supplemental Table 1).

At hospital admission, the two cohorts presented comparable pulmonary dysfunction as assessed by PaO_2_/FiO_2_ ratio (Supplemental Table 1). The Charlson Comorbidity Index was significantly higher (mean 4,3) in the vaccinated group when compared to the unvaccinated one (mean 2,9), indicating more comorbidities among vaccinated than unvaccinated COVID-19 patients (Figure 1A). Remarkably, laboratory tests on hospital admission revealed a significant increase of the levels of serum ferritin as well as of lactate dehydrogenase in the unvaccinated subjects compared to the vaccinated ones (Figure 1B-C). On the contrary, no significant differences were found in the levels of other inflammatory markers (C-reactive protein, IL-6, and D-dimer) between the two groups (Figure 1D-F). Other demographic and clinical features, including treatment and type of vaccine administered, are summarized in Supplemental Table 1.

**Figure 1.**
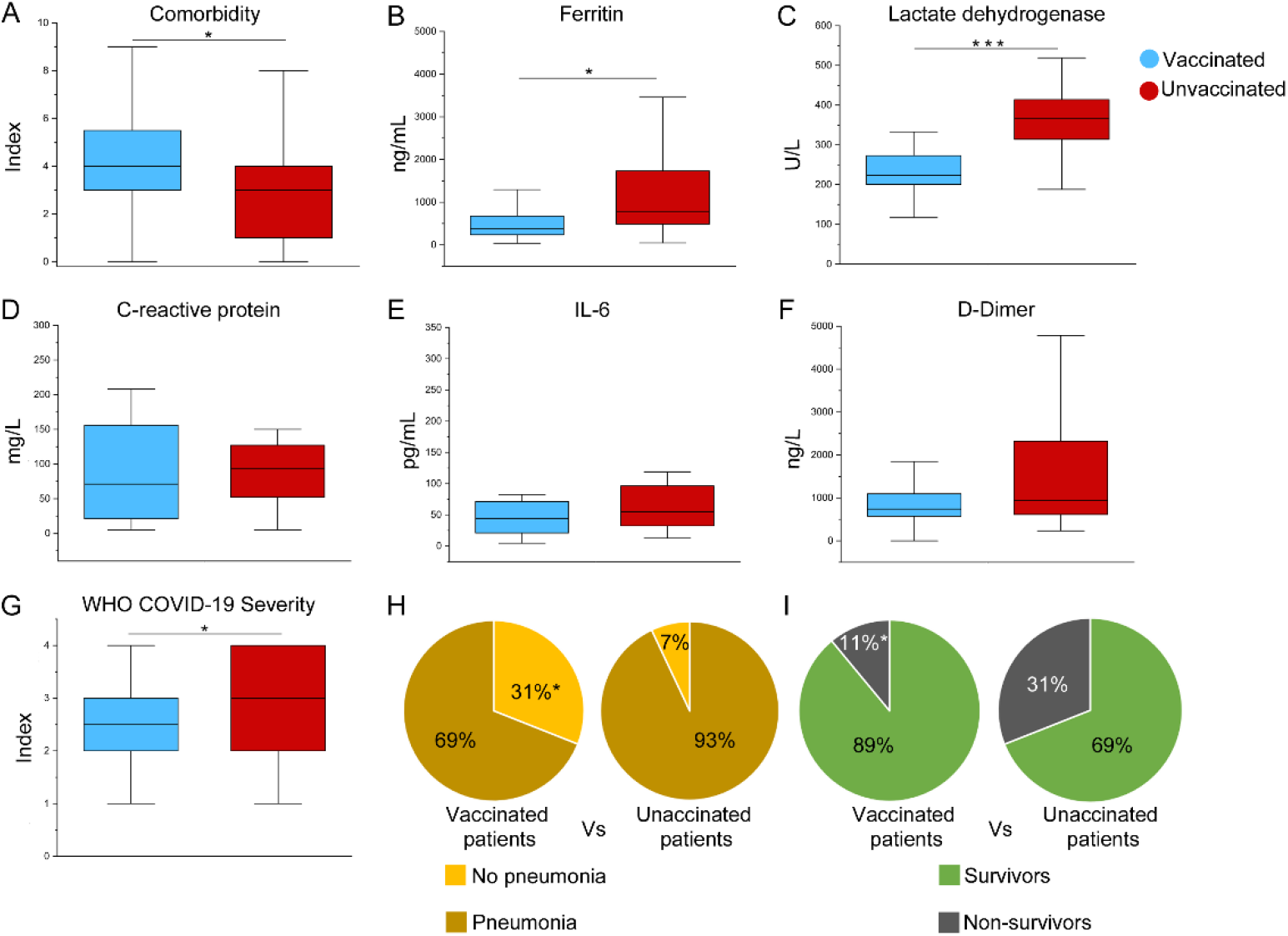
Clinical features of vaccinated and unvaccinated COVID-19 patients requiring hospitalization. Comorbidity index (**A**), serum titers of ferritin (**B**), lactate dehydrogenase (**C**), C-reactive protein (**D**), interleukin-6 (**E**), and D-dimer (**F**) and WHO COVID-19 severity index of 36 vaccinated (blue box) and 29 unvaccinated (red box) hospitalized COVID-19 patients. (**G**). (**H**) Percentages of vaccinated and unvaccinated COVID-19 hospitalized patients who developed pneumonia, on a total of 65 patients. (**I**) Percentages of vaccinated and unvaccinated COVID-19 hospitalized patients who deceased, on a total of 65 patients. Medians and 25^th^ and 75^th^ percentile are shown in boxes. Minimum and maximum values are shown as whiskers. *P<0,05 calculated with Mann-Whitney U test and χ^2^ test.

During hospitalization, significant differences in disease severity were observed between the two groups. According to a scale based on WHO disease severity classification criteria including 0 (asymptomatic), 1 (mild), 2 (moderate), 3 (severe), and 4 (critical), unvaccinated COVID-19 patients presented significantly higher disease severity index than vaccinated patients (Figure 1G). Consistently, 93% (27/29) of the unvaccinated group developed pneumonia, versus 69% (24/36) of the vaccinated patients (Figure 1H). Considering the outcome, mortality rate was 31% (9/29) in the unvaccinated cohort, while 11% (4/36) among the vaccinated group (Figure 1I). All survived subjects were discharged approximately 9 days after hospital admission (Supplemental Table 1). Comprehensively, these data indicated more severe disease in unvaccinated patients compared to vaccinated subjects.

### Immune cell landscape reveals marked alterations in unvaccinated patients

Severe SARS-CoV-2 infection has a profound impact on cells of the immune system. With the aim of producing a general characterization of the immunological profile of vaccinated and unvaccinated hospitalized individuals diagnosed with COVID-19, we evaluated leukocyte subsets’ distribution and features at hospital admission. Despite the two groups were found similar in terms of absolute numbers of white blood cells (WBC), specific subsets displayed significant differences. While neutrophils and basophils presented similar levels in both cohorts, eosinophils, monocytes and lymphocytes’ absolute numbers were significantly reduced in the unvaccinated with respect to the vaccinated group (Figure 2A). Following the same trend, the unvaccinated patients showed decreased absolute numbers of both circulating CD141+ dendritic cells (cDC1 DCs) and CD1c+ (cDC2) DCs, as well as plasmacytoid DCs (pDCs) (Figure 2B). Neutrophil activation markers such as CD11b, CD64 and CD66b were also found to be expressed at similar levels in both groups (Figure 2C). The monocyte compartment was further characterized: CD64 and CD11b expression levels resulted similar between the two groups, while HLA-DR showed a trend towards reduced expression in the unvaccinated cohort (Figure 2C). In addition, we dissected the proportions of classical, intermediate and non-classical subsets. Interestingly, we found significantly lower frequencies of CD14+/CD16++ (non-classical, M3) monocytes in the unvaccinated patients than in vaccinated subjects, while the opposite trend applied for CD14++/CD16- (classical, M1) monocytes, which presented significantly higher frequencies in the unvaccinated group. On the contrary, the CD14++/CD16+ (intermediate, M2) monocyte subset’s frequencies were comparable between the two cohorts (Figure 2D).

**Figure 2.**
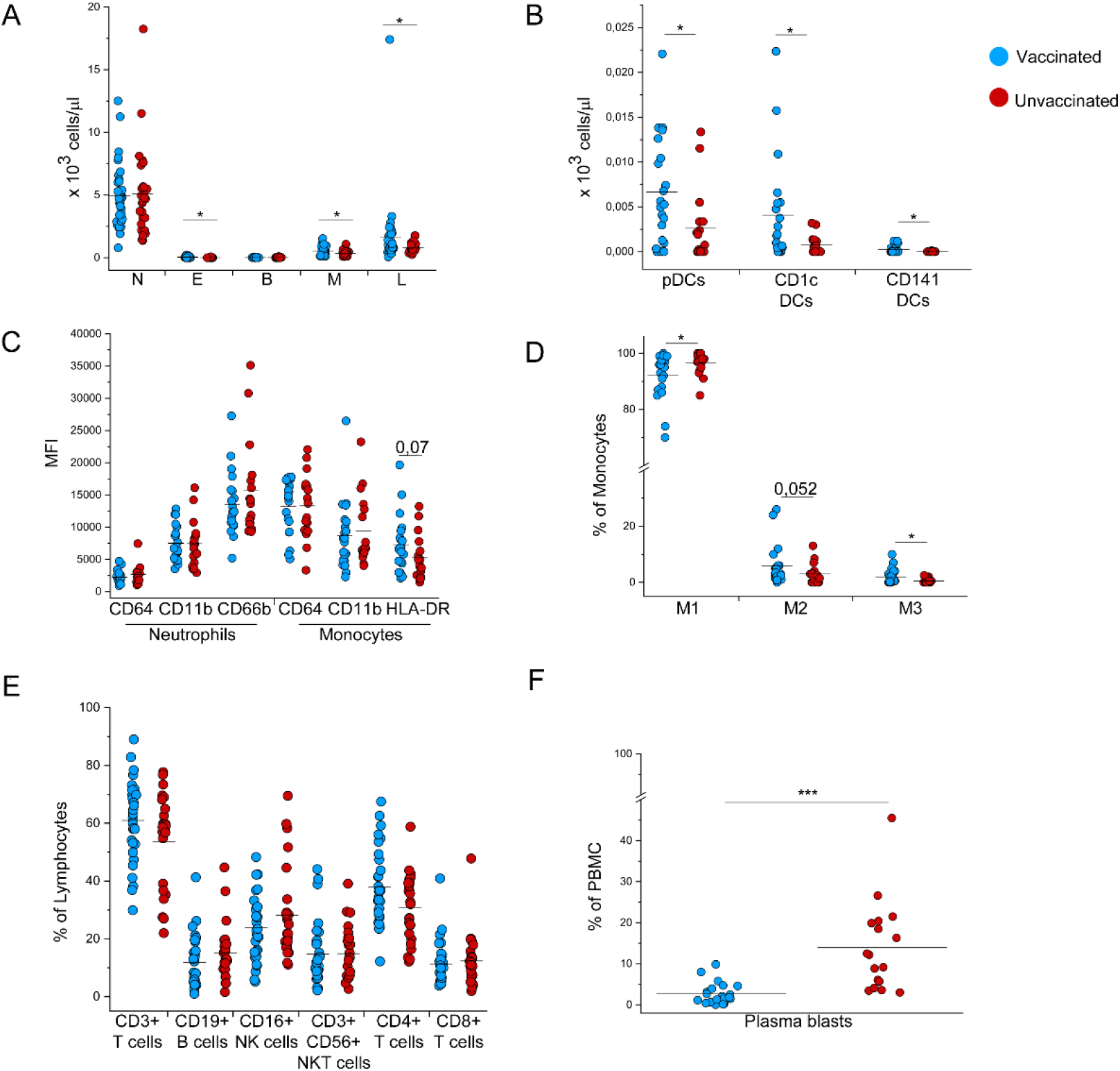
Characterization of circulating immune cell subsets in vaccinated and unvaccinated COVID-19 patients requiring hospitalization. (**A**) Absolute numbers of neutrophils (N), eosinophils (E), basophils (B), monocytes (M) and lymphocytes (L) in 36 vaccinated (blue dots) and 29 unvaccinated (red dots) COVID-19 patients requiring hospitalization. (**B**) Absolute numbers of plasmacytoid dedritic cells (pDCs), CD1c+ dendritic cells and CD141 dendritic cells in 22 vaccinated (blue dots) and 17 unvaccinated (red dots) COVID-19 patients requiring hospitalization. (**C**) Mean Fluorescence Intensity of neutrophil activation markers (CD64, CD11b and CD66b) and monocyte activation markers (CD64, CD11b and HLA-DR) in 22 vaccinated (blue dots) and 17 unvaccinated (red dots) COVID-19 patients requiring hospitalization. **D**) Frequency of CD14++CD16-classical monocytes (M1), CD14+CD16+ intermediate monocytes (M2) and CD14+CD16++ non-classical monocytes (M3) in vaccinated (blue dots) and unvaccinated (red dots) COVID-19 patients requiring hospitalization. (**E**) Frequency of CD3+ T cells, CD19+ B cells, CD16+ NK cells, CD3+CD56+ NKT cells, CD4+ T cells and CD8+ T cells in 30 vaccinated (blue dots) and 26 unvaccinated (red dots) COVID-19 patients. (**F**) Frequency of plasma blasts in vaccinated (blue dots) and unvaccinated (red dots) COVID-19 patients requiring hospitalization. Black lines indicate mean values. *P<0,05, ***P<0,001 calculated with Mann-Whitney U test.

Regarding lymphocyte subsets, no significant differences were detected in the main subpopulations, including CD3+ T cells, CD19+ B cells and CD16+ NK cells. Among CD3+ T cells, the two cohorts exhibited comparable frequencies of CD4+, CD8+ T cells and CD56+ NKT cells (Figure 2E). Further exploring the immune cell composition, we remarkably found a significant increase in the frequency of circulating plasma blasts in unvaccinated compared to vaccinated COVID-19 patients (Figure 2F). Gating strategies for the identification of myeloid and lymphoid cell subsets are reported in Supplemental Figures 2 and 3.

We further characterized CD4+ and CD8+ T cells’ subsets to define naive and memory subpopulations (Supplemental Figure 4). While no differences were detected in the context of CD4+ T cells, the CD8+ naive compartment showed significantly higher frequencies in the unvaccinated than the vaccinated group (Supplemental Figure 5A-B). Similar expression of CXCR3 and CCR6 among CD4+ T cells, as markers of Th1 and Th17 cell polarization, as well as in CXCR5 expression, as marker of T follicular helper cell phenotype was found in both groups (Supplemental Figure 5C-D). Finally, since we and others showed that T cell function is significantly impaired in COVID-19 patients (Mazzoni et al, 2020, Chen & Wherry et al.,2020), we evaluated the basic functionality of the T cell compartment by studying the production of cytokines with anti-viral activity (i.e., IFN-γ, TNF-α and IL-2) after stimulating peripheral blood mononucleated cells with superantigen Staphylococcal Enterotoxin B (SEB). Concerning the CD4+ T cell subset, we also evaluated the expression of activation surface marker CD154. Results of the stimulation showed significantly reduced levels of IL-2 and TNF-α production in the unvaccinated CD4+ T cell compartment, whilst no differences were found in the levels of CD154 expression and IFN-γ production (Supplemental Figure 5E). As for CD8+ T cells, we observed an overall comparable cytokine production between vaccinated and unvaccinated patients (Supplemental Figure 5FB). We also explored T cell polyfunctionality, as T cells are capable of more than one function are superior in terms of antiviral activity (Larsen et al. 2011, Mazzoni et al. 2020). Notably, the percentage of CD4+ T cells exhibiting two to four functions (CD154+, IL-2+, TNF-α, IFN-γ) was significantly reduced in the unvaccinated group, while no differences concerning polyfunctionality were observed in the CD8+ compartment (Supplemental Figure 5G-H).

### Vaccinated patients show a higher Spike-specific humoral and B cell response than unvaccinated patients

It is conceivable that SARS-CoV-2 breakthrough infections requiring hospitalization may be the result of suboptimal response to vaccination or waning immunity. For this reason, we tested humoral and cellular anti-SARS-CoV-2 immunity in vaccinated and unvaccinated COVID-19 patients at hospital admission. Regarding antibody levels, while anti-N IgG and anti-S IgM titers resulted comparable in the two groups (Figure 3A-B), the levels of anti-S IgGs and neutralizing Ig were strikingly higher in the vaccinated than in the unvaccinated subjects (Figure 3C-D). In agreement with this result, flow cytometric analysis of S-specific B cells showed significantly higher frequencies in the vaccinated COVID-19 patients’ group than the unvaccinated counterpart (Figure 3E-F). Evaluation of surface markers CD38 and CD27 coupled with BCR S-specific binding was used to study the frequencies of S-specific plasma blasts, which did not display significant differences between the two groups of patients (Figure 3G-H). Moving to the characterization of T cell response to SARS-CoV-2, we identified by flow cytometry CD4+ T cells reactive to S or M and N by stimulating patients’ PBMCs with specific peptide pools. Reactive T cells were defined on the basis of CD154 expression and production of at least one cytokine between IL-2, TNF-α and IFN-γ. Notably, our results showed that the S-specific and M plus N-specific CD4+ T cell response was comparable between the two patients’ groups (Figure 3I-L).

**Figure 3.**
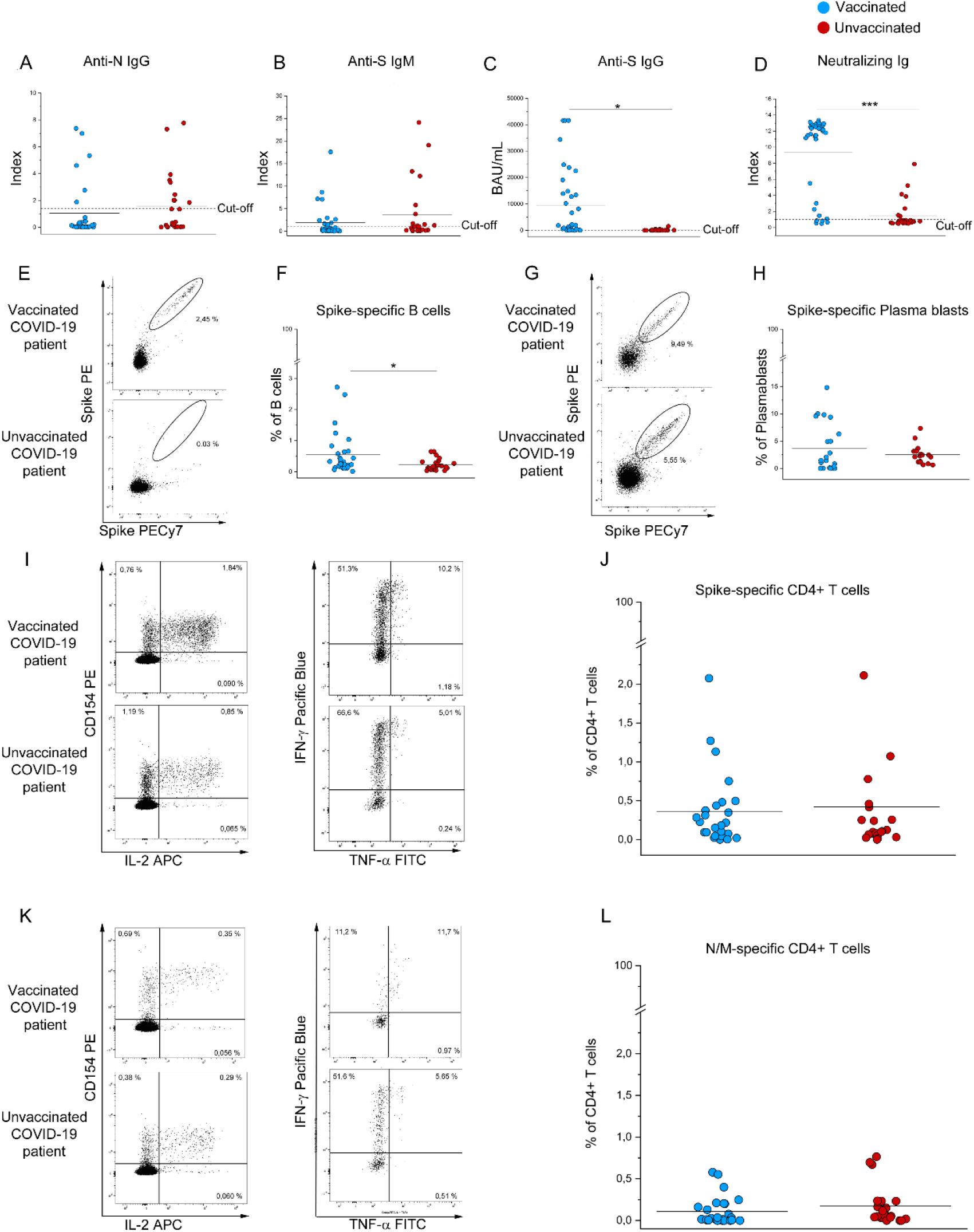
Evaluation of SARS-CoV2-specific humoral response and circulating CD4+ T cells in vaccinated and unvaccinated COVID-19 patients requiring hospitalization. Anti-N IgG (**A**), Anti-S IgM (**B**), Anti-S IgG (**C**) and neutralizing Ig (**D**) titers in vaccinated (blue dots) and unvaccinated (red dots) COVID-19 patients requiring hospitalization. (**E-F**) Representative plots and frequency of Spike-specific B cells in 30 vaccinated (blue dots) and 26 unvaccinated (red dots) COVID-19 patients and Spike-specific plasma blasts (**G**-**H**) in 22 vaccinated (blue dots) and 18 unvaccinated (red dots) COVID-19 patients requiring hospitalization. Representative plots and frequency of Spike-specific (**I**-**J**) and SARS-CoV-2 Nucleoprotein and Membrane (**K-L**) CD4+ T cells defined by CD154+ expression and production of at least 1 cytokine among IL-2, IFN-γ, and TNF-α in 26 vaccinated (blue dots) and 22 unvaccinated (red dots) COVID-19 patients requiring hospitalization. Black lines indicate mean values. *P<0,05; ***P<0,001 calculated with Mann-Whitney U test.

### Deceased patients show reduced anti-Spike immunity at hospital admission

In order to find possible predictive values of patients’ outcome, we compared anti-SARS-CoV-2 immunity at hospital admission between survivors (52/65) and non-survivors (13/65), regardless of vaccination status. Anti-N IgGs and anti-S IgMs did not differ between the two groups (Figure 4A-B), while anti-S IgGs and neutralizing Ig were significantly higher in the group of survivors (Figure 4C-D). As for S-specific B cells, no statistically significant differences were observed between the two cohorts (Figure 4E). Regarding T cell response, CD4+ T cells reactive to N or M showed comparable frequencies in survivors and non-survivors, while S-specific CD4+ T cells showed a significantly lower frequency in the latter (Figure 4F-G). When considering only vaccinated patients, among the 4 non-survivors 2 were older than the average of the whole group (72 years). (Figure 5A). Notably, the youngest vaccinated non-survivor (V19) displayed a higher comorbidity index than the average of the group (Figure 5B). Non-survivors V8, V19 and V28 displayed reduced levels of anti-Spike IgGs and neutralizing Ig than the complete vaccinated cohort (Figure 5C-D). Finally, V8, V28 and V36 exhibited a lower S-specific CD4+ T cell response than the average of all vaccinees (Figure 5E).

**Figure 4.**
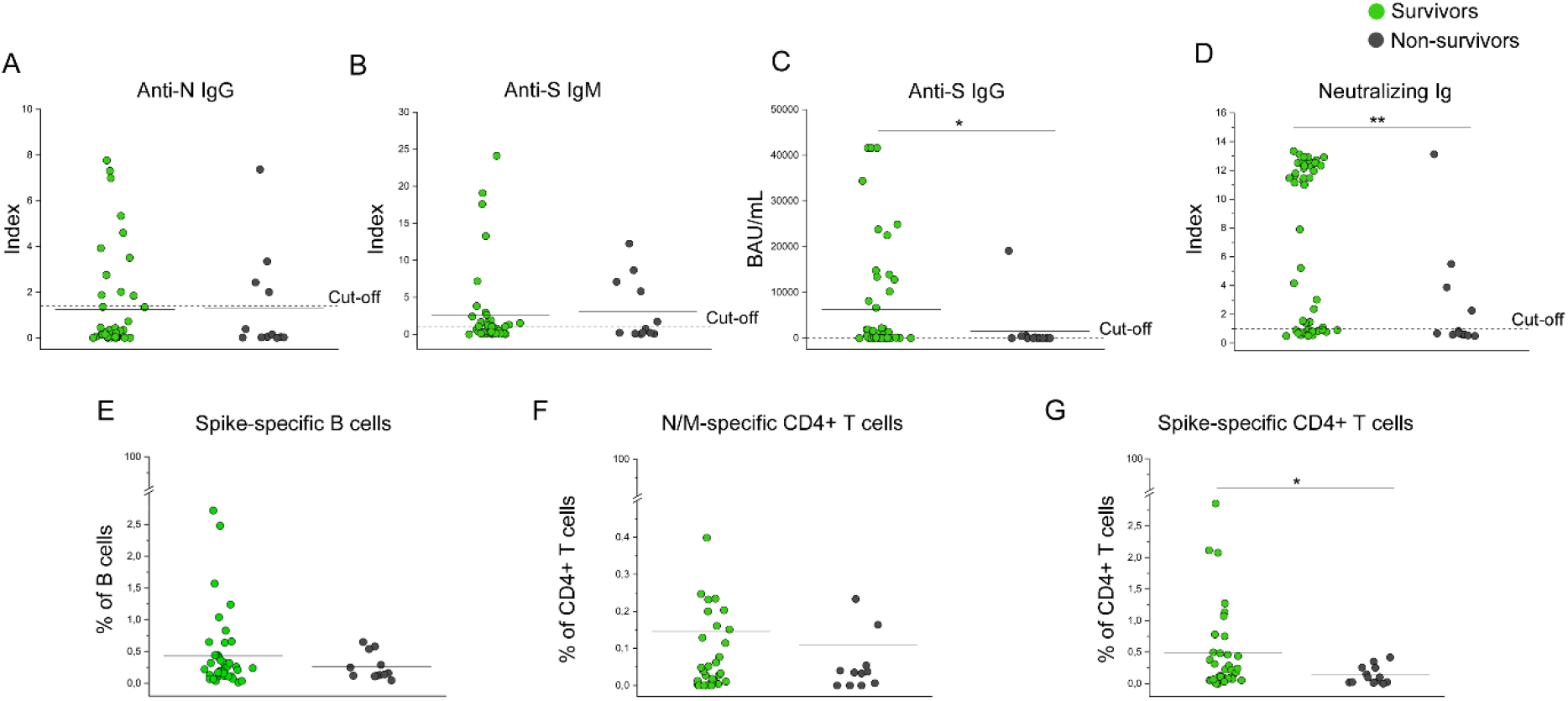
Assessment of SARS-CoV2 specific humoral and cellular response in survived and deceased COVID-19 patients. (**A**) Titers of anti-Nucleoprotein IgGs, anti-Spike IgMs (**B**) and anti-Spike IgGs (**C**) in (n=52) survivors (green dots) and (n=13) non-survivors (dark gray dots) among COVID-19 hospitalized patients.(**D**) Frequency of Spike-specific B cells in 44 survivors (green dots) and 12 non-survivors (dark gray dots) among COVID-19 hospitalized patients. (**E**) Frequency of Nucleoprontein and Membrane specific CD4+ T cells in 35 survivors (green dots) and 12 non-survivors (dark gray dots) among COVID-19 hospitalized patients. (**F**) Frequency of Spike-specific CD4+ T cells in 35 survivors (green dots) and 13 non-survivors (dark gray dots) among COVID-19 hospitalized patients. Black lines indicate mean values. *P<0,05; **P<0,01 calculated with Mann-Whitney U test

**Figure 5.**
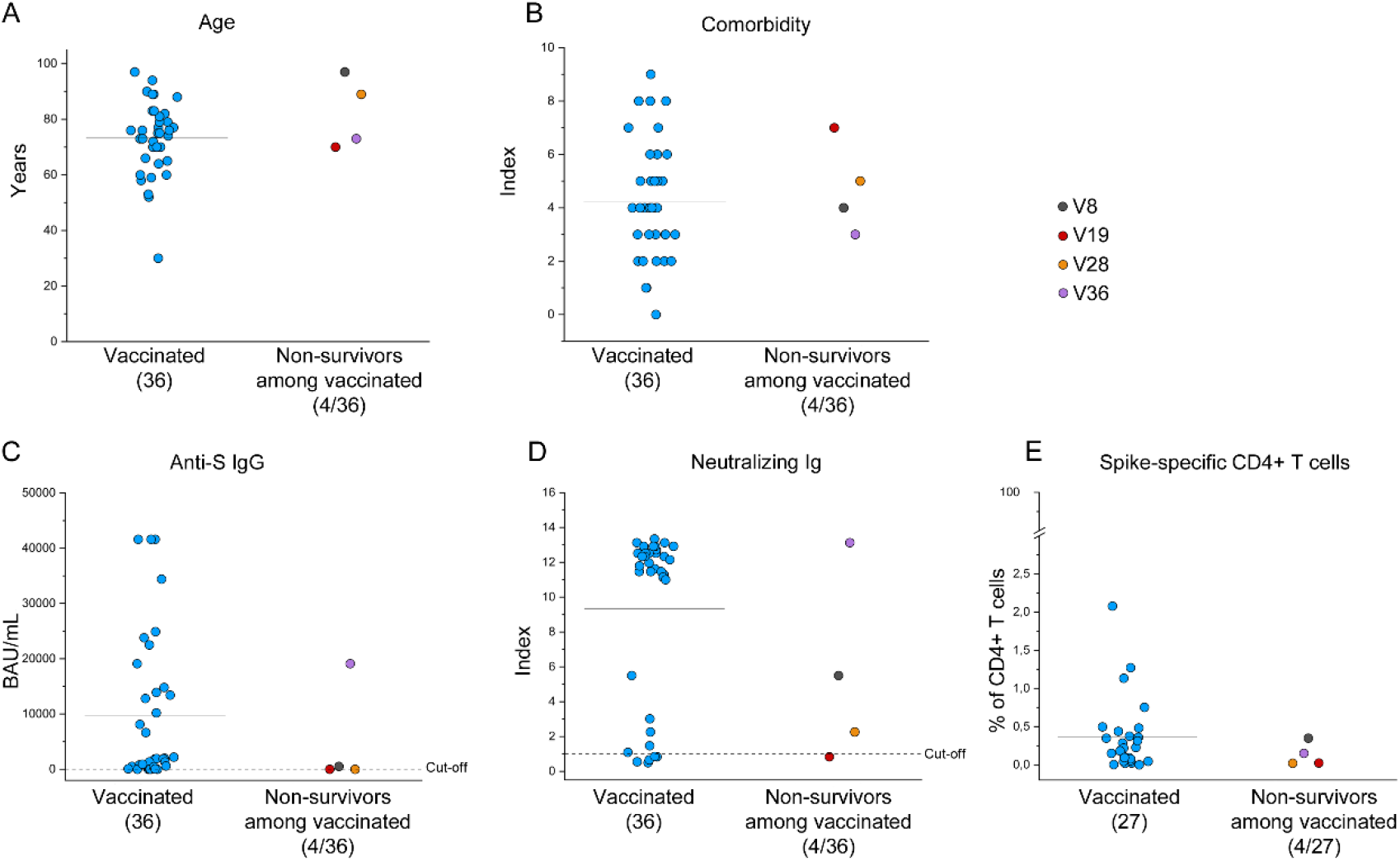
Clinical features and Spike-specific immune response of non-survivors among vaccinated patients. Mean age (**A**) and comorbidity index (**B**) of the 36 vaccinated patients (blue dots) and 4 non-survivors among the vaccinated group (multicolor dots). (**C-D**) Anti-S IgG and neutralizing Ig titres of vaccinated patients (bluedots) and non-survivors among the vaccinated group (multicolor dots). (**E**) Frequency of Spike-specific CD154+ producing at least 1 cytokine among IL-2, IFN-γ, and TNF-α among CD4+ T cells in 27 vaccinated patients (bluedots) and 4 non-survivors among the vaccinated group (multicolor dots). Black lines indicate mean values.

#### Vaccination can prevent fatal outcome in patients with high anti-IFN-α autoantibodies

Since it has been demonstrated that autoantibodies targeting type I IFNs predispose to severe COVID-19 development (Bastard et al., 2020, Zhang et al., 2022), we measured the concentration of anti-IFN-α antibodies in all the study participants. As a control, we enrolled 15 young (<40 years), sex-matched, healthy subjects. We found that 6 out of 65 patients (9,2%) exhibited high anti-IFN-α autoantibodies titers (Figure 6A). Notably, 5 of these 6 patients were male, confirming previous observations showing that anti-type I IFNs antibodies occur more frequently in males (Bastard et al., 2020). Mean age of the 6 patients with high anti-IFN-α antibodies was 79 years, while the rest of the cohort displayed a mean age of 70 years (*p*=0.07, Mann-Whitney U test). Of note, the only female subject among the 6 with high anti-IFN-α autoantibodies was the eldest. Regarding the vaccination status, anti-IFN-α autoantibodies-positive patients were equally divided between the vaccinated and the unvaccinated groups (3/36, 8% vs 3/29, 10% respectively) (Figure 6B). Remarkably, taking into account the outcome, all vaccinated patients with autoantibodies targeting IFN-α were discharged, while all those unvaccinated deceased (Figure 6B).

**Figure 6.**
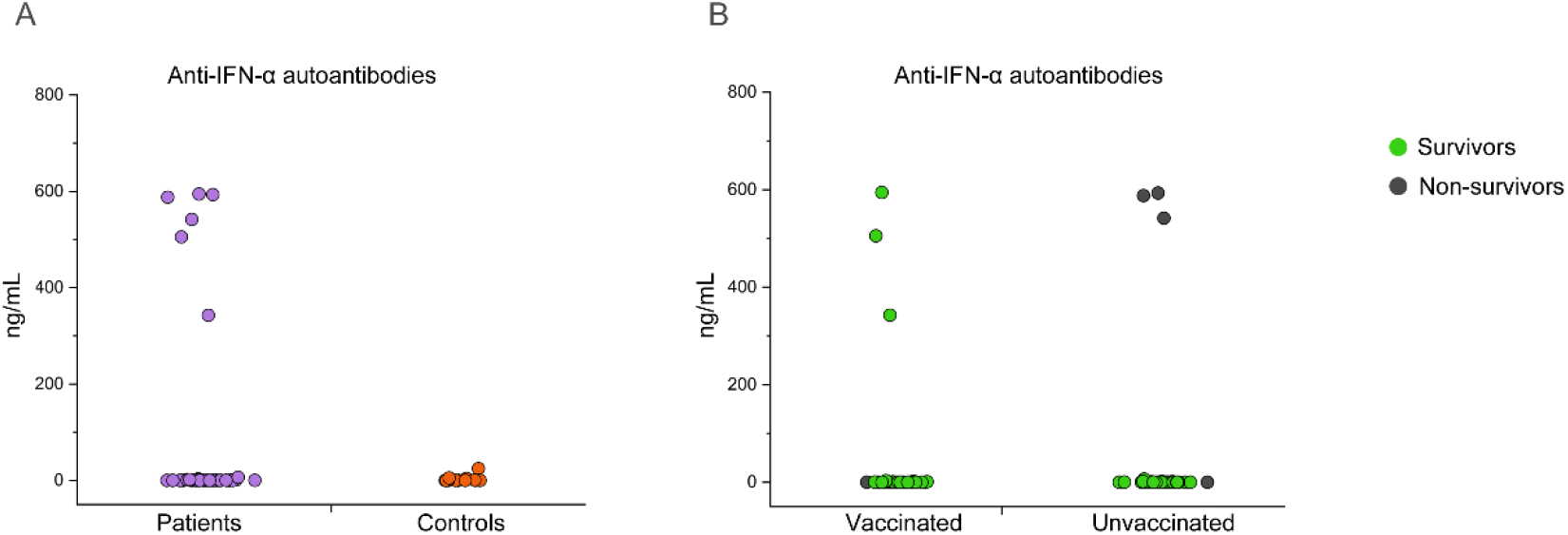
Detection of anti-IFN-α antibodies in COVID-19 patients and controls. Titers of autoantibodies targeting IFN-α in all COVID-19 patients (65) participating to the study (violet dots) and control healthy donors (15) (orange dots). (**B**) Titers of autoantibodies targeting IFN-α in vaccinated (36) and unvaccinated (29) COVID-19 patients, marked basing on positive outcome (green dots) and negative outcome (dark gray dots).

## Discussion

SARS-CoV-2 vaccines represented a turning point in the COVID-19 pandemic, however as we move forward with global vaccination campaigns many unanswered questions need to be addressed. Clinical trials and real-world studies reported high vaccine efficacy against both disease and infection (Zheng et al., 2022). Nonetheless, the progressive waning of antibody levels over time, together with the emergence of new viral variants with increased transmissibility and immune escape potential, led to increased breakthrough infections (Juthani et al., 2021, Kuhlmann et al., 2022). This phenomenon has been associated to a significant surge in the numbers of vaccinated, SARS-CoV-2 infected subjects requiring hospitalization (Arbel et al., 2021 Juthani et al., 2021), but it is currently unknown whether disease progression in hospitalized vaccinated patients mirrors that of unvaccinated patients or differences exist between the two groups. For this reason, we performed a head-to-head comparison of clinical, laboratory and immunological features of vaccinated and unvaccinated COVID-19 patients, who were prospectively enrolled between November and mid-December 2021, during the Delta wave in Italy (ECDC Dashboard). Our data showed that the cohort of vaccinated subjects displayed more risk factors for severe COVID-19 development than unvaccinated patients, including older age and more comorbidities. At hospital admission, despite comparable respiratory function, unvaccinated patients showed significantly higher levels of ferritin and LDH. Since the first descriptions of COVID-19 cases, high serum ferritin levels were associated with severe and critical disease and finally deemed as a prognostic marker of COVID-19 (Wu et al. 2020, Zhou et al. 2020, Para et al. 2021, Qeadan et al. 2021). Moreover, the cell-death marker LDH is a hallmark of severe COVID-19 (Henry et al., 2020), and has been correlated to respiratory failure and ARDS development (Poggiali et al., 2020). The two cohorts displayed similar viral loads, suggesting that initial viral replication is not restrained by previous immunization. The disease course was also remarkably different between the two groups, as the unvaccinated cohort showed a higher frequency of patients developing pneumonia, with an overall higher COVID-19 disease severity score. More importantly, we observed a significantly higher rate of non-survivors in the unvaccinated than the vaccinated group. Altogether, our data suggest that, despite being older and affected by more comorbidities, vaccinated subjects have a substantially more favorable disease course than unvaccinated subjects.

Previous studies have shown that severe COVID-19 has a profound impact on the immune system (Mazzoni et al., 2020, De Biasi et al., 2020). For this reason, in this study we also investigated the main features of innate and adaptive immunity in our cohorts. Unvaccinated patients displayed reduced absolute numbers of circulating eosinophils, monocytes and lymphocytes, as well as reduced numbers of CD1c+ and CD141+ myeloid DC as well as of pDCs. These observations are consistent with previous findings showing that in severe COVID-19 there is selective depletion of these cell subsets (Mazzoni et al., 2020, Peruzzi et al., 2020). Indeed, lymphopenia is one of the hallmarks reflecting disease severity (Mazzoni et al 2020). Focusing on monocyte composition, we observed that unvaccinated patients had a lower frequency of circulating inflammatory (non-classical, M3) monocytes, with a parallel increase in the classical (M1) subset. Comparably, this pattern of monocyte redistribution has been previously demonstrated in severe patients (Peruzzi et al. 2020), and has been associated to the selective transmigration of non-classical monocytes to inflamed lungs (Sanchez-Cerillo et al., 2020). Regarding the lymphoid compartment, we did not find major abnormalities in the main populations of circulating cells, nor in the composition of naïve and memory T cell subsets. Notably, we found significantly higher frequencies of circulating plasma blasts in the unvaccinated cohort. Massive egress of plasma blasts in the circulation has been previously described in the context of other acute infections, including Dengue, Hepatitis A (HAV), influenza and respiratory syncytial virus (Wrammert et al., 2012, Hong et al., 2013, Lee et al., 2011). This observation is consistent with data from another group (De Biasi et al., 2020) and suggests that also in the context of SARS-CoV-2 infection, there is a massive release of antibody secreting cells early after the infection, and it correlates with disease severity. It is currently debated if plasma blasts response is driven mainly by antigen-specific cells, or if bystander cells are also involved (Hong et al., 2013Lee et al., 2011). Our data showing that only a minor fraction of total circulating plasma blasts is S-specific are in agreement with those observed in the context of HAV infection, although obtained with a different experimental approach. However, it should be noted that we monitored the specificity towards only one viral antigen, thus underestimating the total SARS-CoV-2-specific plasma blasts response.

Severe COVID-19 patients are characterized by an altered T cell functionality, with reduced production of cytokines with anti-viral activity (Mazzoni 2020, Chen 2020). This impairment can be the result of an excessive stimulation by pro-inflammatory cytokines, although it remains to be formally proven. In agreement with the increased disease severity of unvaccinated patients, CD4+ T cells from these patients showed reduced production of IL-2 and TNF-α, and reduced percentages of polyfunctional CD4+ T cells than the vaccinated counterpart after stimulation with a superantigen. Regarding SARS-CoV-2-specific immunity, we found that anti-S IgGs and S-specific B cells were significantly higher in vaccinated patients, confirming that hospitalization can also occur in patients that responded to the vaccine. Anti-S IgMs and anti-N IgGs were comparable between the two cohorts, and below cut-off values in most of the patients. This finding is in agreement with the recent and comparable onset of the disease in the two groups. Moreover, this observation further supports that the difference in the levels of anti-S B cells and IgGs in the two cohorts are related to vaccination. Surprisingly, both S- and N plus M-specific CD4+ T cells showed comparable frequencies in the two cohorts. It should be noted T cells were isolated the day of patients’ hospital admission, approximately 9 days after the onset of symptoms. Therefore, it is likely that priming and reactivation of SARS-CoV2 specific T cells, respectively in the unvaccinated and the vaccinated group, produced a comparable result in the two cohorts at this time point from the infection. In agreement, it has previously been demonstrated that a rapid T cell response occurs in patients with favorable outcome (Tan et al., 2021). For this reason, we also recast the criteria to divide our cohort and compared survivors versus non-survivors, regardless of their vaccination status. Anti-S IgGs were significantly higher in survivors, as well as S-specific B cells, despite not reaching statistical significance. Importantly, the frequency of S-specific CD4+ T cells was significantly higher in survivors versus non-survivors. Of note, when considering only vaccinated patients, we found that pre-existing conditions including age and comorbidities, together with suboptimal vaccine response, may contribute to the worst outcome in this group. Previous data have demonstrated that an impaired type I IFN response, due to inborn errors or autoantibodies, is a risk factor for severe COVID-19 occurrence (Bastard et al 2020 and 2021, Zhang et al., 2022). In our study, 9.2% of COVID-19 hospitalized patients exhibited high anti-IFNα levels. Consistently with previous observations, anti-IFN-α autoantibodies-positive patients were predominantly males and in older age groups. Here, we found that among COVID-19 hospitalized patients with autoantibodies targeting IFN-α, those who were vaccinated survived, while the unvaccinated died. Although our data were obtained on a relatively small cohort, this observation further strengthens the concept that patients with anti-type I IFNs antibodies are at risk to develop severe COVID-19. Nonetheless, vaccination can substantially protect even this subset of patients.

In conclusion, our data showed that vaccinated hospitalized COVID-19 patients, most likely infected with the Delta variant, displayed a better disease progression than unvaccinated patients in spite of the unfavorable pre-existing clinical conditions. These clinical observations obtained on unvaccinated patients were confirmed by an immunological landscape that reflected the one previously described in severe COVID-19 patients.

Collectively, these data confirm that a rapid activation of anti-SARS-CoV-2 immunity is mandatory to guarantee patients’ survival. This observation univocally couples a favorable outcome with vaccination, which can even overcome pre-existing risk factors. Indeed, a prior immunization provides a significant advantage in the race to limit the path towards disease worsening.

## Materials and Methods

### Patients

Sixty-five COVID-19 patients were enrolled at the Careggi University Hospital (Azienda Ospedaliero-Universitaria Careggi), Florence, Italy, by the Infective and Tropical Diseases Unit. Among these, 36 patients were vaccinated with any approved viral vector or mRNA SARS-CoV-2 vaccine and had eventually received a booster dose, while 29 were unvaccinated. SARS-CoV-2 infection was confirmed by routine diagnostic PCR amplification of viral genes from nasopharyngeal swabs. Peripheral blood (PB) was collected at hospital admission in EDTA tubes for mononuclear cells (PBMC) recovery, or in clot-activator tubes for serum collection. The clinical and demographic details of each patient can be found in Supplemental Table 1.

### Evaluation of serum clinical parameters

Evaluation of WBC count was performed using a XN 550 haematology analyser (Sysmex Corporation, Kobe, Japan). Serum concentration of C-reactive protein (CRP), ferritin, lactate dehydrogenase (LDH) and interleukin-6 (IL-6) was performed in a Cobas analyser (Roche Diagnostics, Penzberg, Germany). D-dimer and fibrinogen were measured in a ACLTOP550 system (Instrumentation Laboratory, Werfen Group, Kirchheim bei Munchen, Germany). All these parameters were evaluated on blood specimens collected at the same time of those for flow cytometric analysis.

### Myeloid cells immunophenotyping

A stain-lyse-and-then-wash procedure was used to stain surface markers using 100 μl of whole blood. Antibodies used for flow cytometry analysis are listed in Supplemental Table 2. Data acquisition was performed using a 3-laser, 8-colour flow cytometer (FACSCanto TMII, BD Biosciences, San Jose, CA) and then analysed by Infinicyt software (Cytognos SL, Salamanca, Spain). 10^5^ total leucocytes were acquired for each analysis.

### Lymphoid cells immunophenotyping by flow cytometry

PBMCs were obtained after density gradient centrifugation of blood samples. After 2 washes in PBS, cells were stained for 15 minutes with fluorochrome-conjugated monoclonal antibodies (mAbs). Full list of all fluorochrome-conjugated mAbs is in Supplemental Tables S3 and S4. Samples were acquired on a BD LSR II flow cytometer (BD Biosciences) and analysed with FlowJo v10 software. All flow cytometric analyses were performed following published guidelines (Cossarizza et al., 2021).

### Evaluation of SARS-CoV-2-specific IgM and IgG

Evaluation of anti-Spike protein (in trimeric form) IgG (Diasorin); anti-Spike protein IgM (Abbot), anti-Nucleoprotein IgG (Abbott), neutralizing antibodies which block binding of Spike protein with the ACE2 receptorAb (Dia.Pro Diagnostic Bioprobes) was performed following manufacturers’s instructions. The antibody reactivity of each specimen was expressed in BAU/ml, or by the ratio between optical density and cut-off value (index).

### Spike Specific B cells and Plasma blasts evaluation by flow cytometry

For spike-specific B cells and plasma blasts evaluation, 2 million PBMNCs were stained for 30 minutes at 4°C with fluorochrome-conjugated antibodies listed in Supplemental Tables 5 and 6. Recombinant biotinylated SARS-Cov2 Spike protein (Miltenyi Biotech) was conjugated separately with streptavidin PE and PE-Vio770 for 15 minutes at room temperature and pooled in 1:2 ratio, before being added to final staining mix. After the incubation, cells were washed with PBS/EDTA buffer (PEB), and incubated 5 minutes with 7-AAD for viability evaluation. Samples were acquired on a BD LSR II flow cytometer (BD Biosciences). Full gating strategy is reported in Supplemental Figure 6.

### Evaluation of polyclonal- and SARS-CoV-2-specific T cell cytokine production by flow cytometry

For T cells stimulation in vitro, 1.5 million PBMCs, were cultured in complete RPMI plus 5% human AB serum in 96 well flat bottom plates. Cultures were performed in medium alone (background, negative control), with a pool of Spike SARS-CoV-2 peptide pools (Prot_S1, Prot_S+ and Prot_S to achieve a complete sequence coverage of the Spike protein), or with a pool of peptide pools covering nucleoprotein and membrane protein. Each peptide pool was used at 0.6 μM/peptide, accordingly to manufacturer’s instructions (Miltenyi Biotech). In addition, staphylococcal enterotoxin B SEB 1 μg/ml (Sigma Aldrich) was used for the evaluation of polyclonal cytokine production ability. After 2 hours of incubation at 37°C, 5% CO2, Brefeldin A (5 μg/mL) was added, followed by additional 4 hours incubation. Finally, cells were fixed and stained using fluorochrome-conjugated antibodies listed in Supplemental Table 7. Samples were acquired on a BD LSR II flow cytometer (BD Biosciences). Full gating strategy is reported in Supplemental Figure 7. Flow cytometry experiments were performed using published guidelines (Cossarizza et al., 2021)

### Measurement of Human Anti-IFN-α antibodies

Titres of anti-IFN-α antibodies were measured via enzyme-linked immunosorbent assay (Invitrogen) on patients’ sera, according to manufacturer’s instructions. Sera samples of 15 young (<40 years), sex-matched, healthy unvaccinated patients were used as negative controls.

### Statistics

Unpaired, non-parametric, Mann Whitney’s U test and a χ2 test were used for comparison of clinical laboratory findings and for flow cytometric analysis of vaccinated versus unvaccinated COVID-19 patients. P values equal or less than 0.05 were considered significant.

### Study approval

The procedures followed in the study were approved by the Careggi University Hospital Ethical Committee. Written informed consent was obtained from recruited patients.

## Data Availability

All data produced in the present work are contained in the manuscript

## Acknowledgments

We thank all the subjects who participated to the study. This study was supported by funds to the Department of Experimental and Clinical Medicine, University of Florence (Project Excellence Departments 2018-2022), by the University of Florence, project RICTD2122, by the Italian Ministry of Health (COVID-2020-12371849) and by Tuscany Region (TagSARS CoV 2).

