## Supplemental data for "Clinical and immunological features of SARS-CoV-2 breakthrough infections in vaccinated individuals requiring hospitalization"

| | Age, years (range) | Gender | Comorbidity Index | Severity WHO | Pneumonia (yes/no) | $PaO_2/FiO_2$ (mmHg) | Drug therapy | Outcome | Time from onset of symptoms to hospital admission, days | Time from hospital admission to discharge home, days | White blood cell count (x10^9/L) | Neutrophil count (x10^9/L) | Lymphocyte count (x10^9/L) | Monocyte count (x10^9/L) | Eosinophil count (x10^9/L) | Basophil count (x10^9/L) | Red blood cell count (x10^12/L) | Platelet count (x10^9/L) | Lactate dehydrogenase (U/L) | D-dimer (ng/mL) | Serum ferritin (ng/mL) | C-reactive protein (mg/L) | IL-6 (pg/mL) | Anti-IFN-γ antibodies (ng/mL) | Type of vaccine (first+second+third dose) |
|---|---|---|---|---|---|---|---|---|---|---|---|---|---|---|---|---|---|---|---|---|---|---|---|---|---|
| **Reference Range** | - | - | - | 0-4 | - | 400-500 | - | - | - | - | 4-10 | 1,5-7,5 | 0,5-5 | 0,3-1,2 | 0-0,7 | 0-0,2 | 4,2-5,4 | 140-400 | 135-214 | <500 | 15-150 | <5 | <7 | - | - |
| **Vaccinated COVID-19 patients** | | | | | | | | | | | | | | | | | | | | | | | | | |
| V1 | 75-79 | M | 4 | 2 | yes | 295 | S | H | 6 | 6 | 15,5 | 12,51 | 1,74 | 1,16 | 0,05 | 0,05 | 4,54 | 234 | 273 | 602 | 227 | 191 | 303 | 505,5 | A+A |
| V2 | 65-69 | F | 3 | 0 | no | 404 | - | H | - | 9 | 5,19 | 2,35 | 2,39 | 0,31 | 0,12 | 0,02 | 4,26 | 325 | 140 | - | 95 | 8 | - | 0 | P+P |
| V3 | 55-59 | F | 8 | 0 | no | 314 | - | H | - | 14 | 7,91 | 6,57 | 0,65 | 0,59 | 0,08 | 0,02 | 3,3 | 376 | 175 | 1316 | 381 | 71 | - | 1,9 | P+P |
| V4 | 75-79 | M | 7 | 2 | yes | 309 | S, R | H | 6 | 11 | 8,64 | 4,83 | 3,3 | 0,5 | 0 | 0,01 | 4,1 | 186 | 445 | 770 | 1295 | 15 | 19,9 | 0 | P+P |
| V5 | 80-84 | M | 5 | 3 | yes | 271 | S | H | 6 | 16 | 4,03 | 2,62 | 1,14 | 0,25 | 0,01 | 0,01 | 3,6 | 223 | 226 | 866 | 619 | 65 | 32,6 | 0 | P+P |
| V6 | 75-79 | M | 3 | 2 | yes | 283 | S | H | 10 | 6 | 8,24 | 6,86 | 0,65 | 0,59 | 0,11 | 0,03 | 3,49 | 148 | 281 | 739 | 1208 | 179 | - | 342,7 | A+A |
| V7 | 90-94 | F | 4 | 3 | yes | 223 | S | H | 6 | 25 | 5,18 | 4,35 | 0,71 | 0,11 | 0 | 0,01 | 3,98 | 279 | 272 | 1141 | 591 | 202 | - | 594,4 | P+P |
| V8 | 95-99 | F | 4 | 3 | yes | 168 | S | D | 11 | - | 6,6 | 6,01 | 0,44 | 0,14 | 0 | 0,01 | 3,71 | 442 | 265 | 940 | 2390 | 191 | 227 | 0 | P+P+P |
| V9 | 75-79 | F | 3 | 3 | yes | 285 | S, R | H | 7 | 5 | 3,4 | 1,92 | 1,17 | 0,29 | 0,01 | 0,01 | 5,02 | 174 | 240 | 1037 | 37 | 38 | 23,2 | 0 | A+A |
| V10 | 75-79 | F | 5 | 2 | no | 352 | R | H | 3 | 5 | 4,94 | 3,93 | 0,76 | 0,21 | 0,03 | 0,01 | 3,86 | 133 | 213 | 1585 | 220 | 71 | 82,5 | 0 | P+P |
| V11 | 75-79 | M | 4 | 3 | yes | 257 | S | H | 5 | 5 | 8,66 | 7,79 | 0,5 | 0,36 | 0 | 0,01 | 3,57 | 134 | 223 | 1184 | 355 | 182 | - | 0 | N+N |
| V12 | 60-64 | M | 2 | 3 | yes | 309 | S | H | 7 | 6 | 5,49 | 2,88 | 2,15 | 0,39 | 0,06 | 0,01 | 5,5 | 136 | 278 | 642 | 332 | 36 | 41,8 | 0 | A+A |
| V13 | 55-59 | M | 3 | 3 | yes | 219 | S, R, M | H | 4 | 15 | 20,7 | 2,46 | 17,41 | 0,81 | 0 | 0,02 | 4,63 | 149 | 283 | 389 | 709 | 86 | - | 0 | N+N |
| V14 | 90-94 | F | 4 | 2 | no | - | S | H | 14 | 3 | 6,81 | 4,89 | 1,16 | 0,73 | 0,02 | 0,01 | 3,81 | 212 | 210 | 1247 | 442 | 156 | 71,2 | 0 | P+P |
| V15 | 80-84 | F | 6 | 4 | yes | 233 | S | H | 10 | 24 | 9,54 | 8,45 | 0,69 | 0,38 | 0 | 0,02 | 4,34 | 182 | 265 | 611 | 1038 | 35 | 46,2 | 0,5 | P+P |
| V16 | 70-74 | M | 8 | 0 | no | - | M | H | - | 9 | 6,56 | 4,39 | 1,28 | 0,61 | 0,22 | 0,06 | 4,45 | 266 | 200 | 747 | 258 | 5 | - | 1,0 | P+P+P |
| V17 | 70-74 | M | 3 | 2 | no | 338 | S | H | 9 | 5 | 4,89 | 4,13 | 0,057 | 0,18 | 0 | 0,01 | 4,84 | 250 | 201 | 292 | 237 | 49 | 43,3 | 0 | A+A |
| V18 | 85-89 | F | 9 | 3 | yes | 280 | S, T | H | 3 | 10 | 6,78 | 5,13 | 0,77 | 0,81 | 0,05 | 0,02 | 3,7 | 172 | 235 | 1849 | 216 | 23 | - | 0 | P+P |
| V19 | 70-74 | F | 7 | 4 | yes | 233 | S, R | D | 6 | - | 5,6 | 5,15 | 0,33 | 0,11 | 0 | 0,01 | 3,38 | 111 | 333 | 474 | 265 | 208 | 230 | 0 | P+P |
| V20 | 75-79 | M | 4 | 3 | yes | 252 | S, R | H | 4 | 8 | 4,08 | 2,45 | 1,03 | 0,5 | 0,09 | 0,01 | 5,61 | 118 | 177 | 1093 | 983 | 28 | - | 0 | A+A |
| V21 | 80-84 | M | 6 | 2 | yes | 319 | S, R | H | 4 | 6 | 6,75 | 4,77 | 1,04 | 0,85 | 0,09 | 0,01 | 4,63 | 149 | 194 | 0 | - | 103 | 57,5 | 0 | P+P+P |
| V22 | 50-54 | M | 1 | 4 | yes | 271 | S, T | H | 6 | 16 | 5,91 | 5,15 | 0,63 | 0,13 | 0 | 0 | 4,82 | 147 | 310 | 229 | 888 | 158 | - | 0 | A+A |
| V23 | 75-79 | M | 2 | 2 | yes | 371 | S | H | 10 | 3 | 4,28 | 2,91 | 0,82 | 0,54 | 0 | 0,01 | 4,79 | 208 | 213 | 580 | 507 | 34 | 64,5 | 0 | A+P |
| V24 | 85-89 | F | 8 | 3 | yes | 271 | S, M | H | 6 | 4 | 4,13 | 3,09 | 0,72 | 0,29 | 0,01 | 0,02 | 3,64 | 108 | 201 | 739 | 343 | 82 | 44,9 | 0,7 | P+P+P |
| V25 | 70-74 | M | 5 | 1 | no | 258 | S, M | H | - | 8 | - | - | - | - | - | - | - | - | - | - | - | - | - | 0 | A+A |
| V26 | 60-64 | M | 2 | 3 | yes | 202 | S | H | 9 | 8 | 8,99 | 7,97 | 0,77 | 0,24 | 0 | 0,01 | 5 | 215 | 286 | 499 | 1364 | 172 | - | 0 | J |
| V27 | 75-79 | M | 6 | 0 | no | - | R, M | H | - | 8 | 9,25 | 6,23 | 1,87 | 1,08 | 0,02 | 0,06 | 5,51 | 164 | 233 | 319 | - | 74 | - | 0 | P+P |
| V28 | 85-89 | F | 5 | 3 | yes | 276 | S | D | - | - | 5,9 | 4,74 | 0,62 | 0,51 | 0,02 | 0,01 | 3,23 | 304 | 117 | 3977 | 677 | 111 | - | 0 | P+P |
| V29 | 70-74 | F | 5 | 3 | yes | 276 | S | H | 2 | 5 | 7,51 | 5,4 | 1,41 | 0,51 | 0,18 | 0,01 | 4,13 | 297 | 256 | 1731 | 527 | 89 | - | 0 | A+A |
| V30 | 80-84 | F | 4 | 2 | yes | 390 | - | H | 16 | 3 | 9,91 | 6,09 | 2,79 | 0,9 | 0,07 | 0,05 | 4,83 | 492 | 162 | 1071 | 434 | 21 | - | 0,2 | P+P |
| V31 | 60-64 | M | 2 | 2 | yes | 304 | S, R, M | H | 3 | 5 | 5,26 | 3,41 | 0,86 | 0,96 | 0,01 | 0,02 | 5,08 | 203 | 201 | 624 | 555 | 11 | 5,5 | 3,2 | P+P |
| V32 | 50-54 | M | 1 | 0 | no | 457 | - | H | - | 16 | 14,9 | 11,25 | 1,98 | 1,53 | 0,1 | 0,03 | 3,44 | 274 | 168 | 592 | 154 | 7 | - | 0 | P+P |
| V33 | 70-74 | F | 5 | 3 | yes | 271 | S | H | 10 | 11 | 4,49 | 3,48 | 0,82 | 0,18 | 0 | 0,01 | 4,09 | 173 | 200 | | 177 | 9 | 21,1 | 0 | A+A |
| V34 | 65-69 | F | 2 | 1 | no | - | - | H | 7 | 1 | 4,03 | 0,79 | 2,43 | 0,72 | 0,08 | 0,01 | 4,39 | 147 | 189 | 572 | 209 | 7 | 4,5 | 0 | P+P |
| V35 | 30-34 | M | 0 | 1 | no | 438 | - | H | 2 | 1 | 6,49 | 3,41 | 1,87 | 1,07 | 0,11 | 0,03 | 5,55 | 189 | 202 | 490 | 236 | 10 | 5,4 | 0 | P+P |
| V36 | 70-74 | F | 3 | 4 | yes | 282 | S, T | D | 12 | - | 6,59 | 5,29 | 1,11 | 0,18 | 0 | 0,01 | 4,85 | - | 313 | 612 | 277 | 143 | - | 0 | A+A |
| **Mean** | **73,2** | - | **4,3** | **2,3** | - | **294,1** | - | - | **7,0** | **8,7** | **7,2** | **5,0** | **1,7** | **0,5** | **0,0** | **0,0** | **4,3** | **215,3** | **233,7** | **895,7** | **552,9** | **82,0** | **73,6** | **40,3** | - |
| **SD** | **13,3** | - | **2,2** | **1,2** | - | **65,2** | - | - | **3,5** | **5,9** | **3,6** | **2,5** | **2,8** | **0,4** | **0,1** | **0,0** | **0,7** | **91,6** | **62,2** | **697,8** | **482,6** | **68,8** | **86,9** | **138,1** | - |

**Unvaccinated COVID-19 patients**

| | **Age, years** | Gender | **Comorbidity Index** | **Severity WHO** | **Pneumonia (yes/no)** | $PaO_2/FiO_2$ (mmHg) | Drug therapy | **Outcome (discharged home/deceased)** | Time from onset of symptoms to hospital admission, days | Time from hospital admission to discharge home, days | White blood cell count (x10^9/L) | Neutrophil count (x10^9/L) | **Lymphocyte count (x10^9/L)** | **Monocyte count (x10^9/L)** | **Eosinophil count (x10^9/L)** | Basophil count (x10^9/L) | Red blood cell count (x10^12/L) | Platelet count (x10^9/L) | **Lactate dehydrogenase (U/L)** | D-dimer (ng/mL) | **Serum ferritin (ng/mL)** | C-reactive protein (mg/L) | IL-6 (pg/mL) | Anti-IFN-γ antibodies (ng/mL) | Type of vaccine (first +second +third dose) |
|---|---|---|---|---|---|---|---|---|---|---|---|---|---|---|---|---|---|---|---|---|---|---|---|---|---|
| N1 | 60-64 | F | 2 | 2 | yes | 333 | S | H | 8 | 10 | 3,19 | 2,19 | 0,71 | 0,28 | 0 | 0,1 | 4,33 | 102 | 239 | 413 | 93 | 22 | 17 | 1,7 | na |
| N2 | 70-74 | M | 3 | 2 | yes | 309 | S, R, M | H | 8 | 10 | 8,9 | 7,77 | 0,62 | 0,5 | 0 | 0,01 | 4,09 | 279 | 331 | 734 | 693 | 5 | 12,6 | 0 | na |
| N3 | 60-64 | M | 2 | 4 | yes | 314 | S, R, M | D | 8 | - | 8,26 | 7,38 | 0,5 | 0,37 | 0 | 0,01 | 4,47 | 141 | 399 | 559 | 599 | 107 | 119 | 0,8 | na |
| N4 | 45-49 | F | 0 | 4 | yes | 242 | S, R, T | H | 6 | 12 | 3,72 | 3,2 | 0,38 | 0,14 | 0 | 0 | 4,4 | 146 | 455 | 937 | 167 | 150 | - | 0 | na |
| N5 | 75-79 | M | 8 | 4 | yes | 276 | S, R, M | D | 2 | - | 19,4 | 18,25 | 0,55 | 0,54 | 0 | 0,03 | 5,09 | 343 | 367 | 4786 | 679 | 271 | 243 | 593,3 | na |
| N6 | 65-69 | M | 2 | 3 | yes | 304 | S | H | - | 6 | 5,92 | 4,36 | 1,04 | 0,47 | 0,02 | 0,03 | 4,97 | 285 | 353 | 1193 | 737 | 114 | 97,6 | 0 | na |
| N7 | 50-54 | M | 1 | 2 | yes | 168 | S, R, M | H | 6 | 9 | 2,27 | 1,93 | 0,25 | 0,09 | 0 | 0 | 4,28 | 150 | 471 | 694 | 1024 | 96 | 64 | 0,9 | na |
| N8 | 90-94 | F | 5 | 3 | yes | - | S, R, M | H | 4 | 5 | 3,25 | 2,16 | 0,74 | 0,33 | 0,01 | 0,01 | 3,22 | 156 | 293 | 7118 | 760 | 10 | - | 0 | na |
| N9 | 55-59 | F | 1 | 1 | no | 314 | M | H | 5 | 2 | 3,36 | 1,37 | 1,77 | 0,21 | 0 | 0,01 | 4,58 | 115 | 280 | 432 | 324 | 10 | 20 | 0 | na |
| N10 | 85-89 | M | 6 | 3 | yes | 200 | S | D | 5 | - | 6,85 | 5,68 | 1,06 | 0,1 | 0 | 0,01 | 4,27 | 88 | 519 | 6149 | 4139 | 68 | - | 0 | na |
| N11 | 70-74 | M | 4 | 4 | yes | 252 | S, T, M | D | 12 | - | 12,8 | 11,51 | 0,79 | 0,49 | 0 | 0,01 | 5 | 234 | 406 | 718 | 480 | 102 | 51,4 | 541,6 | na |
| N12 | 50-54 | F | 1 | 1 | no | 352 | S, M | H | 7 | 2 | 5,08 | 2,69 | 1,76 | 0,62 | 0 | 0,01 | 4,74 | 198 | 188 | 233 | 429 | 5 | - | 0,2 | na |
| N13 | 80-84 | M | 6 | 2 | yes | 357 | S, R, M | H | 8 | 5 | 5,81 | 4,87 | 0,54 | 0,39 | 0 | 0,01 | 4,84 | 239 | 377 | 1224 | 1192 | 71 | 96,3 | 1,3 | na |
| N14 | 70-74 | F | 3 | 4 | yes | 261 | S, R | D | 8 | - | 6,12 | 4,62 | 1,19 | 0,31 | 0 | 0 | 3,67 | 150 | 363 | 1170 | 1879 | 52 | 44,1 | 0,5 | na |
| N15 | 45-49 | M | 0 | 4 | yes | 576 | S, R, M | H | 6 | 12 | 6,32 | 5,52 | 0,66 | 0,12 | 0 | 0,02 | 5,51 | 235 | 1149 | 2322 | 1844 | 90 | 41,8 | 0 | na |
| N16 | 70-74 | F | 4 | 3 | yes | 261 | S | H | 7 | 10 | 6,71 | 5,49 | 1,03 | 0,18 | 0 | 0,01 | 3,44 | 200 | 493 | 1434 | 1957 | 70 | 54,9 | 0,2 | na |
| N17 | 65-69 | M | 3 | 2 | yes | 309 | S | H | 7 | 11 | 2,13 | 1,43 | 0,57 | 0,12 | 0 | 0,01 | 4,78 | 216 | 343 | 612 | 784 | 89 | - | 0 | na |
| N18 | 75-79 | M | 3 | 4 | yes | 130 | S | H | - | 29 | 5,24 | 4,52 | 0,49 | 0,22 | 0 | 0,01 | 4,97 | 151 | 472 | 1151 | 1633 | 57 | - | 0 | na |
| N19 | 75-79 | F | 4 | 3 | yes | 238 | S, T | H | 9 | 9 | 4,02 | 3,19 | 0,58 | 0,25 | 0 | 0 | 5,41 | 213 | 395 | 553 | - | 150 | 60,5 | 0 | na |
| N20 | 70-74 | M | 5 | 4 | yes | 266 | S, M | D | 9 | - | 9,13 | 8,11 | 0,52 | 0,42 | 0,06 | 0,02 | 4,67 | 166 | 285 | 934 | - | 128 | - | 587,9 | na |
| N21 | 45-49 | M | 0 | 2 | yes | 352 | S | H | 13 | 2 | 7,18 | 5,11 | 0,96 | 1,08 | 0 | 0,03 | 4,78 | 342 | 236 | 1158 | - | 93 | - | 0 | na |
| N22 | 65-69 | F | 2 | 4 | yes | 117 | S | D | 9 | - | 4,59 | 3,65 | 0,64 | 0,29 | 0 | 0,01 | 4,47 | 172 | 314 | 843 | 3460 | 125 | 32,3 | 0 | na |
| N23 | 90-94 | F | 4 | 4 | yes | 147 | - | D | 11 | - | 8,46 | 7,6 | 0,57 | 0,25 | 0 | 0,04 | 4,86 | 355 | 400 | 7007 | 47 | 38 | - | 0 | na |
| N24 | 50-54 | M | 2 | 3 | yes | 261 | S | H | 14 | 15 | 6,38 | 4,7 | 1,01 | 0,64 | 0,02 | 0,02 | 4,17 | 456 | 511 | 37629 | 1177 | 106 | - | 6,4 | na |
| N25 | 45-49 | M | 3 | 2 | yes | 338 | S, M | D | 4 | - | 4,25 | 3,71 | 0,41 | 0,12 | 0 | 0,01 | 3,05 | 81 | 324 | 526 | 17459 | 127 | 115 | 0 | na |
| N26 | 45-49 | M | 0 | 3 | yes | 295 | S, R, M | H | 4 | 13 | 7,13 | 5,64 | 1,11 | 0,36 | 0 | 0,02 | 4,59 | 117 | 414 | 458 | 1732 | 147 | 44,4 | 0 | na |
| N27 | 65-69 | M | 4 | 3 | yes | 309 | S, R | H | 5 | 5 | 2,68 | 1,75 | 0,67 | 0,22 | 0,02 | 0,02 | 5,04 | 208 | 403 | 4930 | 1173 | 144 | 59,3 | 0 | na |
| N28 | 80-84 | F | 4 | 3 | yes | 290 | S | H | 5 | 7 | 6,92 | 4,95 | 1,35 | 0,49 | 0,08 | 0,05 | 4,54 | 182 | 343 | 128000 | 517 | 128 | 94,4 | 1,7 | na |
| N29 | 55-59 | M | 1 | 3 | yes | 314 | S, M | H | 9 | 5 | 6,14 | 4,79 | 1,02 | 0,33 | 0 | 0 | 6,07 | 194 | 252 | 778 | 487 | 27 | 19,3 | 0,3 | na |
| **Mean** | **66,6** | - | **2,9** | **3,0** | - | **281,6** | - | - | **7,4** | **9,0** | **6,3** | **5,1** | **0,8** | **0,3** | **0,0** | **0,0** | **4,6** | **203,9** | **392,2** | **7403,3** | **1748,7** | **89,7** | **67,7** | **59,9** | - |
| **SD** | **14,3** | - | **2,0** | **0,9** | - | **87,7** | - | - | **2,9** | **6,1** | **3,5** | **3,4** | **0,4** | **0,2** | **0,0** | **0,0** | **0,7** | **87,8** | **168,6** | **24213,5** | **3347,7** | **58,3** | **53,7** | **178** | - |
| ***p*-value** | **0,0469** | | **0,0134** | **0,0184** | **0,0178** | 0,7724 | - | **0,0459** | 0,575 | 0,7265 | 0,2147 | 0,7978 | **0,0121** | **0,026** | **<0,0001** | 0,4438 | 0,1954 | 0,7511 | **<0,0001** | 0,0677 | **0,004** | 0,5173 | 0,484 | 0,1328 | - |

**Supplemental Table 1. Demographic, clinical, and laboratory characteristics of vaccinated (n=36) and unvaccinated COVID-19 (n=29) patients at hospital admission.**

A: ChAdOx1-S/nCoV-19 vaccine; C: Caucasian; CH: Chinese; D: deceased; H: discharged home; J: Ad26.COV2.S vaccine; L: Latin american; M: antiviral monoclonal antibodies; N: mRNA-1273 vaccine; na: not applicable; P: BNT162b2 vaccine; R: remdesivir; S: steroids; T: tocilizumab.

*p* values are calculated by Mann-Whitney U test or $\chi 2$ test, as appropriate.

| Antigen | Fluorochrome | Clone | Company |
|---|---|---|---|
| CD45 | V-500 | 2D1 | BDBioscience |
| CD66b | V-450 | G10F5 | BDBioscience |
| HLA-DR | V-450 | L243 | BDBioscience |
| CD15 | FITC | MMAC | BDBioscience |
| CD14 | FITC | MϕP9 | BDBioscience |
| CD123 | PE | 9F5 | BDBioscience |
| CD64 | PerCP-5.5 | 10.1 | BDBioscience |
| CD13 | PE-Cy7 | L138 | BDBioscience |
| CD33 | PE-Cy7 | P676 | BDBioscience |
| CD11b | APC | D12 | BDBioscience |
| CD141 | APC-H7 | 1A4 | BDBioscience |
| CD16 | APC-H7 | 3G8 | BDBioscience |
| CD1c | PE | AD5-8E7 | Miltenyi |

**Supplemental Table 2**. List of all fluorochrome mAbs used for flow cytometric analysis of neutrophils, monocytes, plasmacytoid and classical dendritic cells.

| Antigen | Fluorochrome | Clone | Company |
|---|---|---|---|
| HLA-DR | FITC | L243 | BDBioscience |
| CD56 | PE | NCAM16.2 | BDBioscience |
| CD16 | PerCP 5.5 | 3G8 | BDBioscience |
| CD4 | PE-Cy7 | SK3 | BDBioscience |
| CD19 | APC | SJ25C1 | BDBioscience |
| CD8 | APC-H7 | SK1 | BDBioscience |
| CD3 | V450 | ICHT1 | BDBioscience |
| CD45 | V450-C | 2D1 | BDBioscience |

**Supplemental Table 3**. List of all fluorochrome mAbs used for flow cytometric analysis of T cell subsets' immunophenotyping.

| Antigen | Fluorochrome | Clone | Company |
| --- | --- | --- | --- |
| CXCR3 | FITC | FAB160F | R&D Systems |
| CXCR5 | PE | FAB190P | R&D Systems |
| CCR7 | V450 | 150503 | BD Horizon |
| CD45RO | APC | 559865 | BD Pharmigen |
| CD3 | PerCP | SK7 | BDBioScience |
| CD8 | APC-Cy7 | SK1 | BDBioScience |
| CCR6 | PE-Cy7 | 130-100-381 | Miltenyi |
| CD4 | eFluor 506 | RPA-T4 | Invitrogen |

**Supplemental Table 4**. List of all fluorochrome mAbs used for flow cytometric analysis of T follicular and memory cells.

| Antigen | Fluorochrome | Clone | Company |
|---|---|---|---|
| CD19 | APC-Vio770 | LT19 | Miltenyi |
| CD27 | VioBright FITC | M-T271 | Miltenyi |
| IgA | VioGreen | IS11-8E10 | Miltenyi |
| IgG | VioBlue | IS11-3B2.2.3 | Miltenyi |
| CD14 | PerCP | TÜK4 | Miltenyi |
| IgM | APC | PJ2-22H3 | Miltenyi |
| CD3 | PerCP | BW264/56 | Miltenyi |
| 7AAD | | | Miltenyi |
| Spike | PE | | Miltenyi |
| Spike | PE-Vio770 | | Miltenyi |

**Supplemental Table 5**. List of all fluorochrome mAbs used for flow cytometric analysis of antigen specific B cells.

| Antigen | Fluorochrome | Clone | Company |
|---|---|---|---|
| CD38 | APC-Cy7 | HB7 | BDBioScience |
| CD19 | APC | SJ25C1 | BDBioScience |
| CD27 | VioBright FITC | M-T271 | Miltenyi |
| CD14 | PerCP | TÜK4 | Miltenyi |
| CD3 | PerCP | BW264/56 | Miltenyi |
| 7-AAD | PerCP | | Miltenyi |
| CD20 | V 450 | L27 | BDBioScience |
| CD45 | eFluor V500 | 2D1 | BDBioScience |
| Spike | PE | | |
| Spike | PECy-7 | | |

**Supplemental Table 6**. List of all fluorochrome mAbs used for flow cytometric analysis of antigen specific Plasma blasts.

| Antigen | Fluorochrome | Clone | Company |
|---|---|---|---|
| TNF-α | FITC | 6401.1111 B | BDBioscience |
| CD154 | PE | TRAP1 | BDBioscience |
| CD3 | PerCP | SK7 | BDBioscience |
| CD4 | PECy-7 | SK3 | Invitrogen |
| CD8 | SB600 | SK1 | eBioscience™ |
| IL-2 | APC | MQ1-17H12 | BDBioscience |
| IFN-γ | Pacific Blue | B27 | BioLegend |
| L/D | Fixable Viability Stain 780 | | BDBioscience |

**Supplemental Table 7**. List of all fluorochrome mAbs used for flow cytometric analysis of antigen specific and polyclonal T cells.

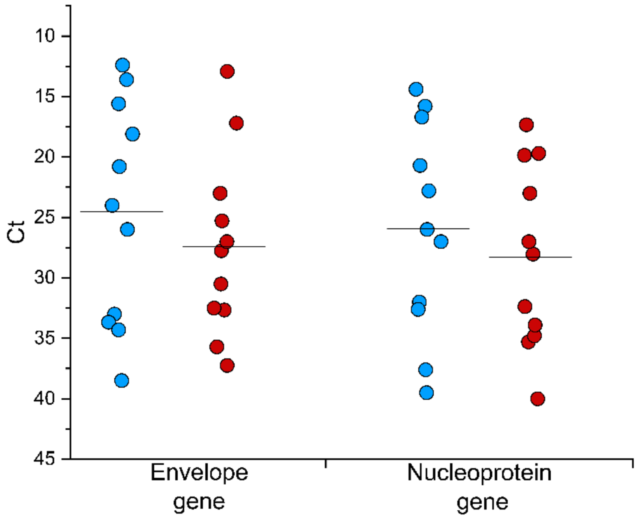

**Supplemental Figure 1. Viral loads of recruited patients**. SARS-CoV2 infection of recruited subjects was confirmed by routine diagnostic PCR amplification of Envelope and Nucleoprotein viral genes from nasopharyngeal swabs. Cycle thresholds (Ct) are shown in blue dots for 11 vaccinated patients and red dots for 11 unvaccinated patients.

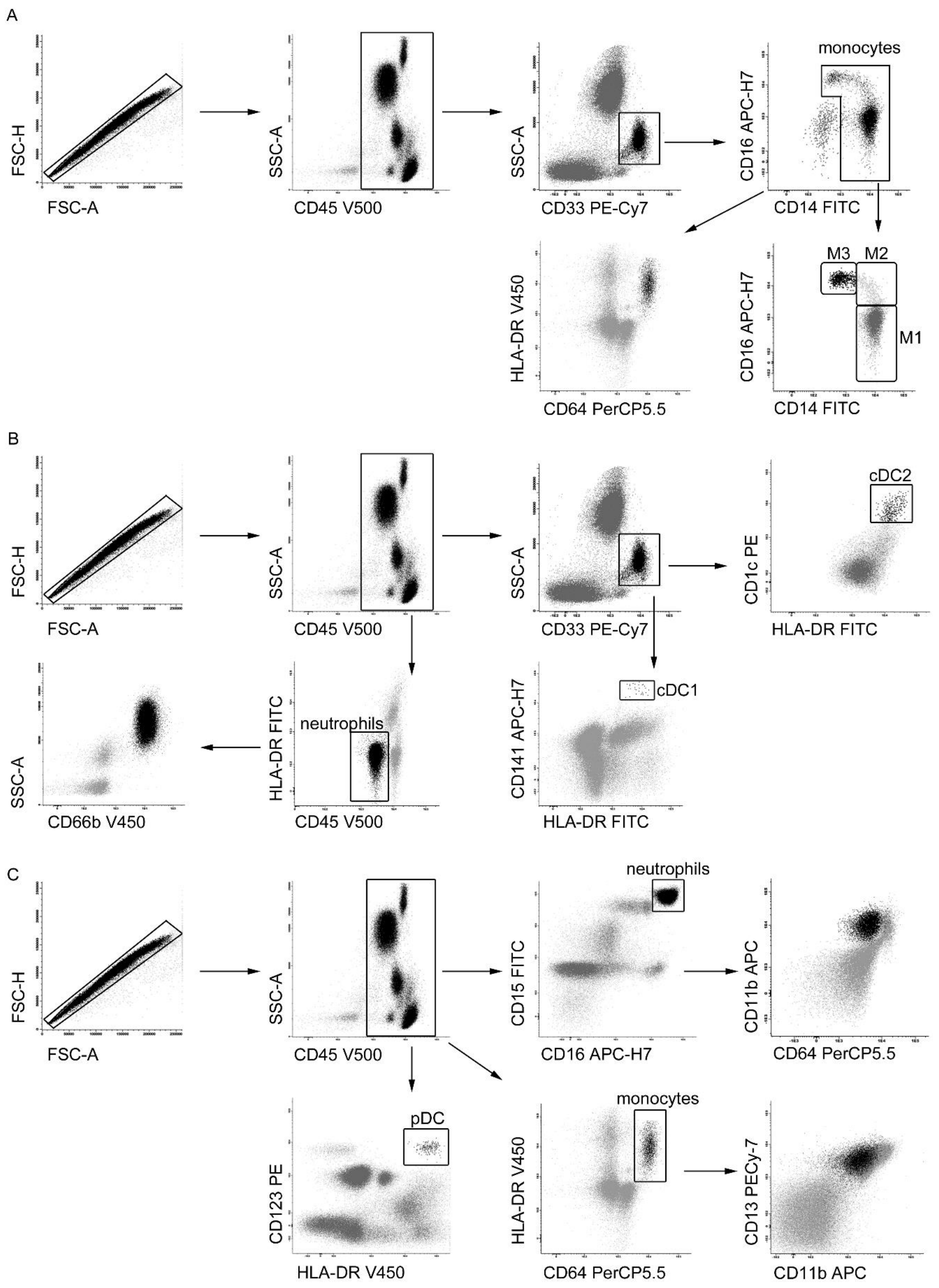

**Supplemental Figure 2. Gating strategy for myeloid cells identification.**

Whole blood cells were gated using physical parameters (FSC, SSC) and CD45 surface marker. (**A**) Monocytes were isolated from CD33+ cells as CD14+CD16+ cells. Then, they were gated basing on the expression of activation markers CD64 and HLA-DR and, in parallel, identified as CD14++(M1), CD14+CD16+ (M2) and CD16++ (M3). (**B**) Dendritic cells were identified through physical parameter

SSC and expression of CD33, then gated into CD1c+ (cDC2) and CD141+ (cDC1). Neutrophils were identified as HLA-DR negative-CD45 dim, and then analyzed for CD66b expression.(**C**) Plasmacytoid Dendritic cells were isolated from CD45+ cells as CD123+ HLA-DR+ cells. Neutrophils were gated from CD45+ cells into CD15++ CD16++ cells and marked by expression of activation markers CD11b, CD64. Monocytes were identified as HLA-DR+ CD64+ and then characterized for CD13 and CD11b expression.

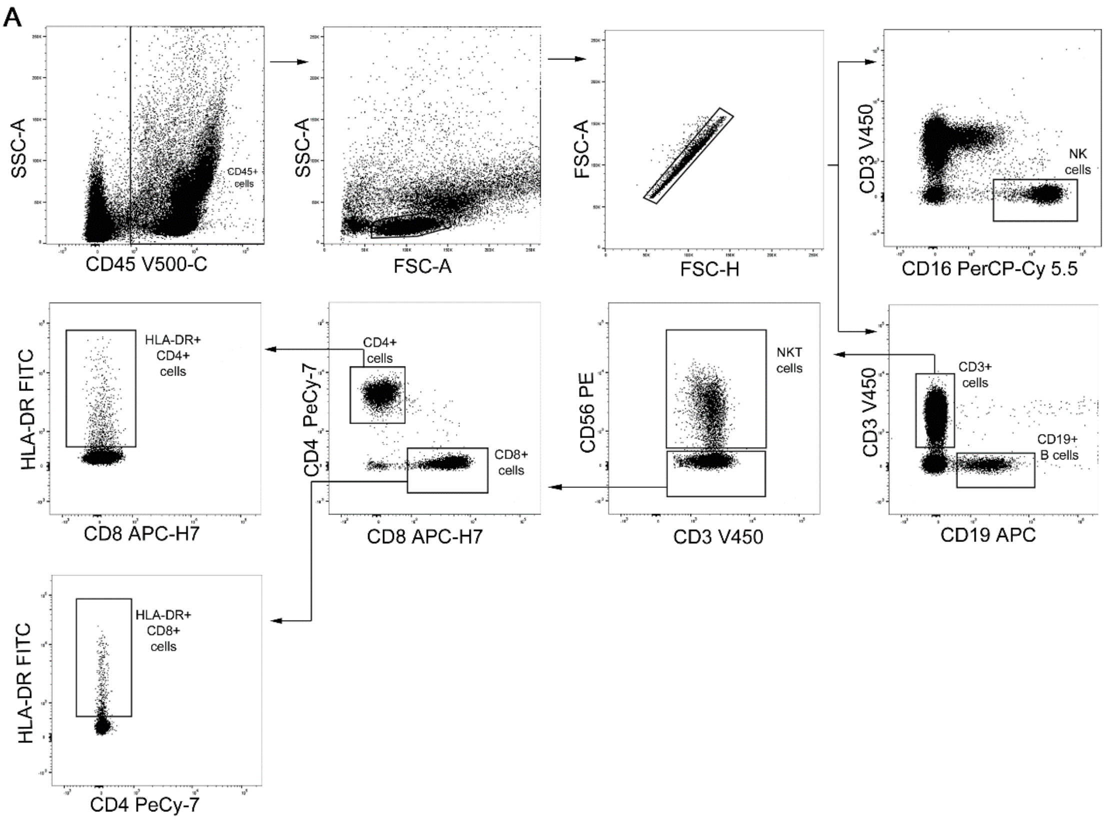

**Supplemental Figure 3. Gating strategy for lymphoid cell subsets immunophenotyping.**

(**A**)PBMCs were gated based on physical parameter SSC and CD45 surface expression, lymphocytes were then gated basing on physical parameters (SSC, FSC). Doublets were removed using FSC-A and FSC-A parameters. NK cells were isolated by CD16 expression. B cells were identified as CD19+ and T cells as CD3+. Within these, CD3+CD56+ cells were identified as NKT. We then identified CD4+ and CD8+ T cells. Among these, we identified HLA-DR expressing cells.

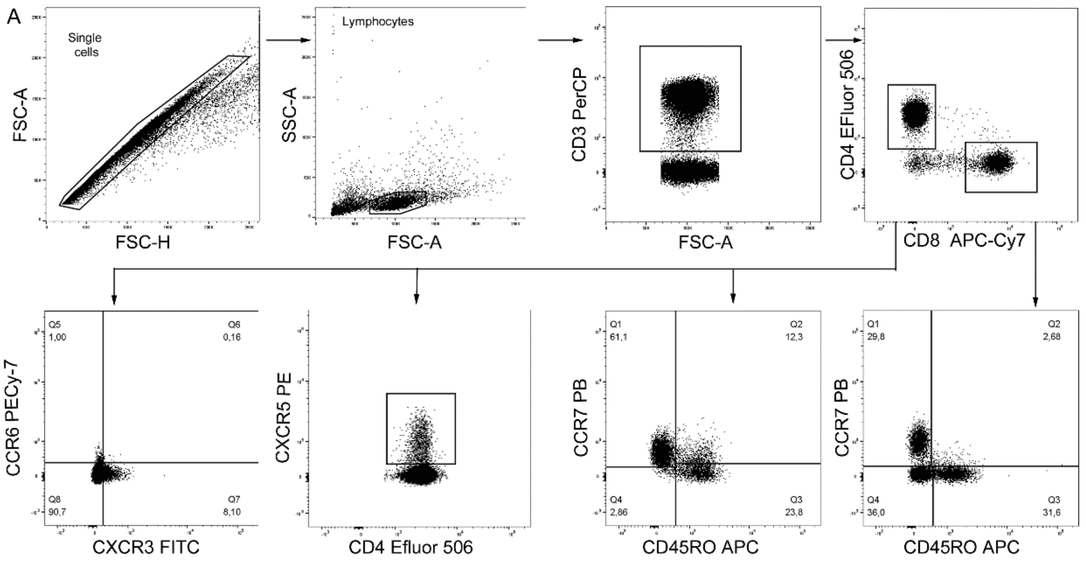

**Supplemental Figure 4. Gating strategy for the identification of naive and memory T cell subsets, and T follicular helper subset.**

(**A**) Doublets were removed using FSC-A and FSC-A parameters and lymphocytes were gated based on physical parameters (FSC-SSC). T cells were identified as CD3+. We then identified CD4+ and CD8+ T cells. Among these, we identified naive (CD45RO-CCR7+), effector memory (CD45RO+CCR7-), central memory (CD45RO+CCR7+), TEMRA (CD45RO-CCR7-). Among CD4+ T cells, we identified CXCR3+CCR6-, CXCR3+CCR6+, CXCR3+CCR6-, CXCR3-CCR6- and CXCR5+ T follicular helper cells.

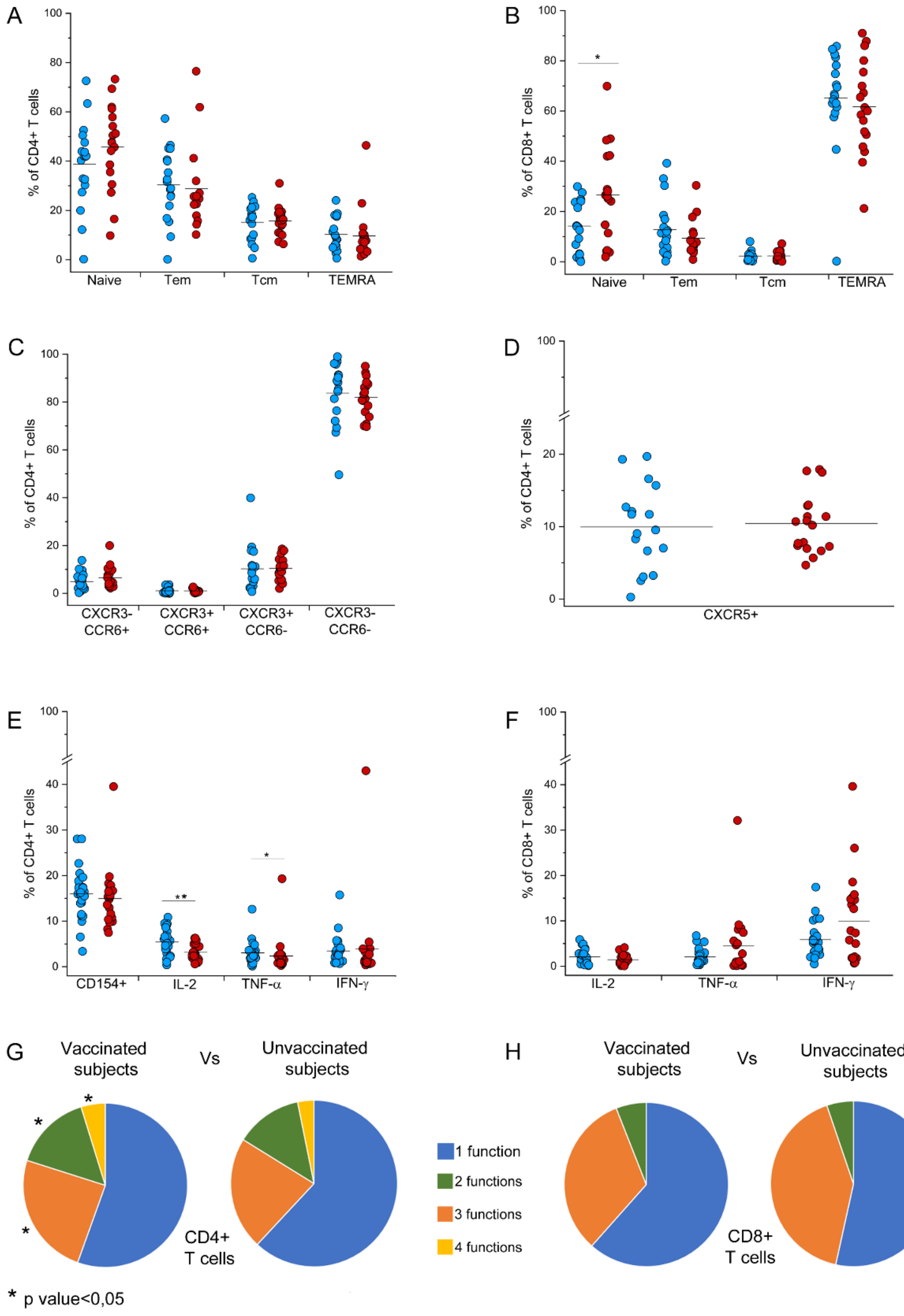

**Supplemental Figure 5. Identification of CD4+ and CD8+ naive and memory T cell subsets, and T follicular helper subset.**

(**A**) Frequency of naive (CD45RO-CCR7+), effector memory (CD45RO+CCR7-), central memory (CD45RO+CCR7+), TEMRA (CD45RO-CCR7-) among CD4+ T cells in 17 vaccinated (blue dots) and 18 unvaccinated (red dots) COVID-19 patients.(**B**) Frequency of naive (CD45RO-CCR7+), effector memory (CD45RO+CCR7-), central memory (CD45RO+CCR7+), TEMRA (CD45RO-CCR7-) among CD8+ T cells in vaccinated (blue dots) and unvaccinated (red dots) COVID-19 patients requiring hospitalization.(**C**) Frequency of CXCR3+CCR6-, CXCR3+CCR6+, CXCR3+CCR6-,CXCR3-CCR6- among CD4+ T cells in vaccinated (blue dots) and unvaccinated (red dots) COVID-19 patients requiring hospitalization.(**D**) Frequency of CXCR5+ T follicular helper cells among CD4+ T cells in vaccinated (blue dots) and unvaccinated (red dots) COVID-19 patients. (**E**) Frequency of CD154+, IL-2, TNF-α and IFN-γ producing cells among CD4+ T cells in vaccinated (blue dots) and unvaccinated (red dots) COVID-19 patients requiring hospitalization. (**F**) Frequency of IL-2, TNF-α and IFN-γ producing cells among CD8+ T cells in vaccinated (blue dots) and unvaccinated (red dots) COVID-19 patients requiring hospitalization. (**G**) Characterization CD4+ T cell polyfunctionality in vaccinated and unvaccinated COVID-19 patients. Results are shown as mean percentages from 26 vaccinated and 22 unvaccinated COVID-19 patients requiring hospitalization. (**H**) Characterization CD8+ T cell polyfunctionality in vaccinated and unvaccinated COVID-19 patients. Results are shown as mean percentages from 26 vaccinated and 22 unvaccinated COVID-19 patients requiring hospitalization. *P<0,05 **P>0,005 ***P<0,0005 calculated with Mann-Whitney U test.

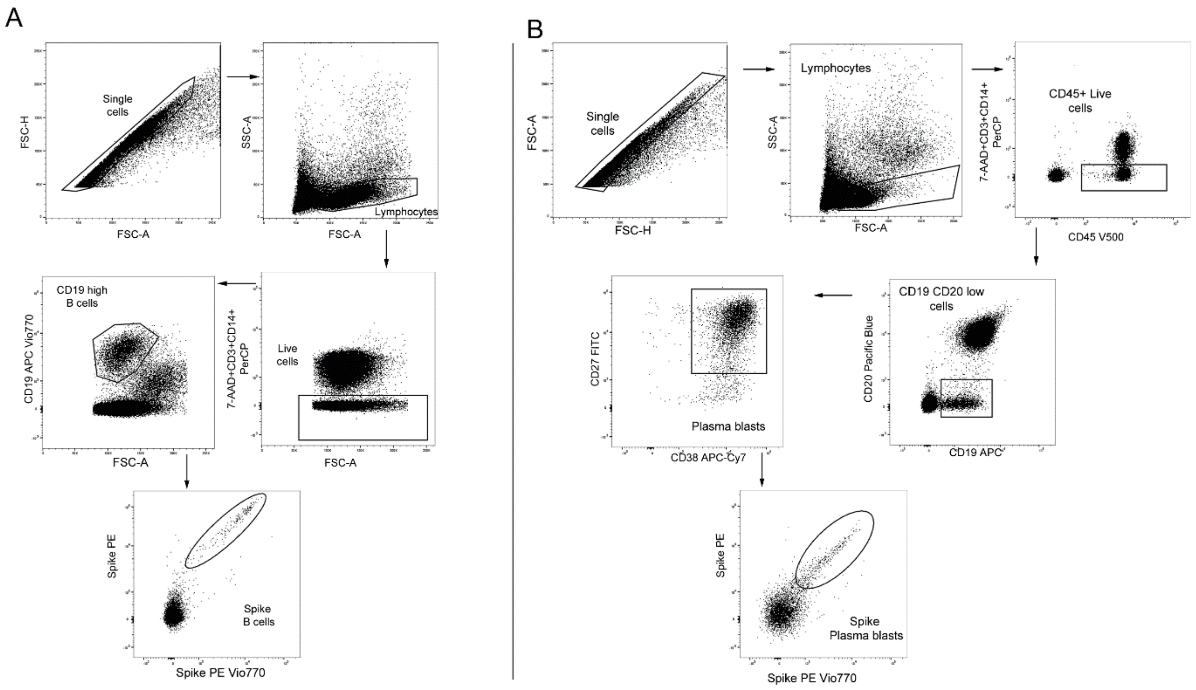

**Supplemental Figure 6. Gating strategy for the identification of Spike specific B cells (A) and Plasma blasts (B).** (**A**) Doublets were removed using FSC-A and FSC-A parameters and lymphocytes were gated based on physical parameters (FSC-SSC). PerCP was used as dump channel for the exclusion of dead cells (7AAD), CD3+ T cells and CD14 monocytes. B cells were identified as CD19+ high. B cells binding PE- and PE Vio770-conjugated spike protein were then identified as spike-specific. (**B**) Doublets were removed using FSC-A and FSC-A parameters and lymphocytes were gated based on physical parameters (FSC-SSC). PerCP was used as dump channel for the exclusion of dead cells (7AAD), CD3+ T cells and CD14 monocytes. Plasma blasts were first identified as CD19+ low, then further gated into CD27+CD38+ cells. Plasma blasts binding PE- and PE Vio770-conjugated spike protein were then identified as spike-specific.

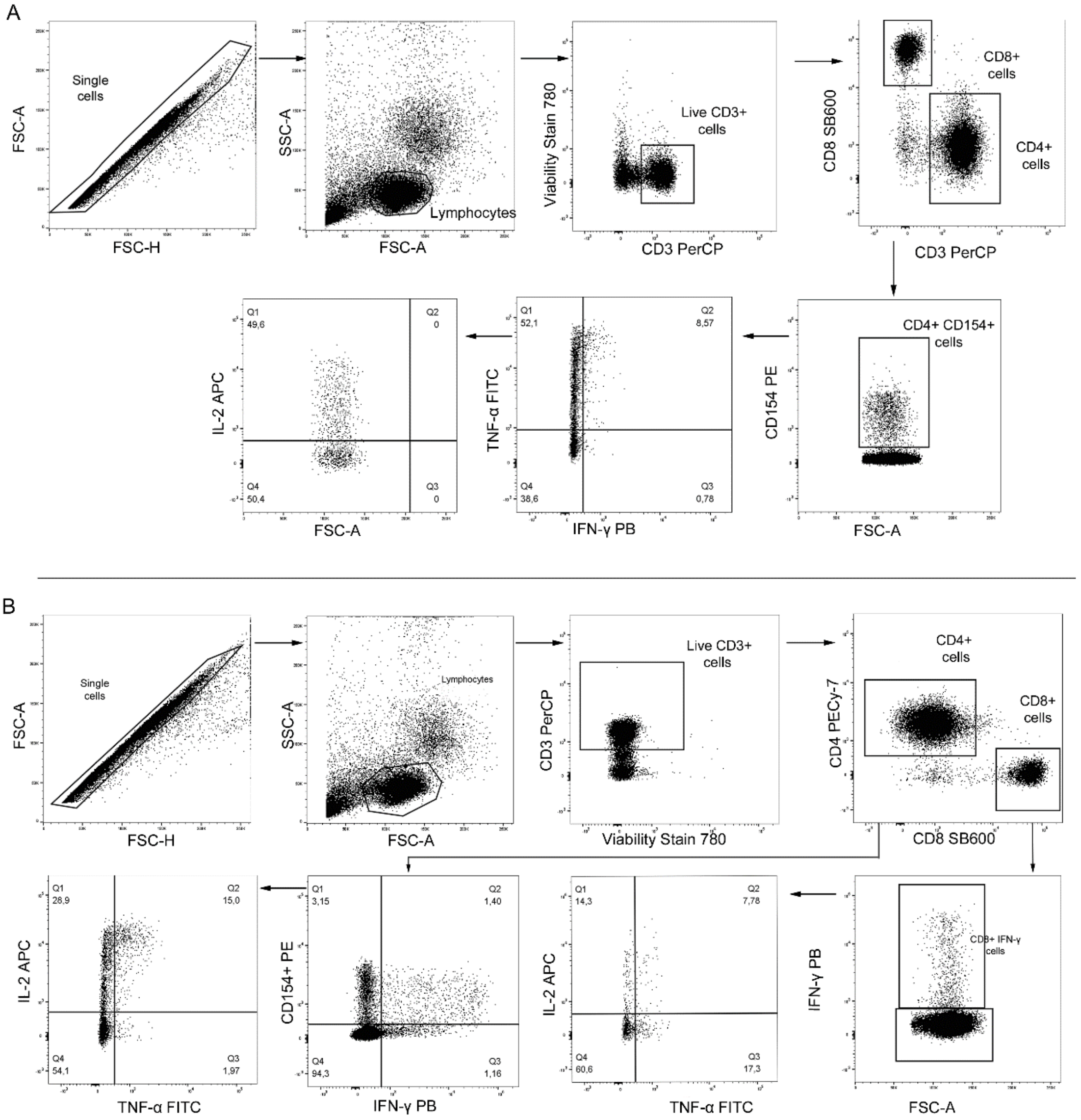

**Supplemental Figure 7. Gating strategy for the identification of polyclonal (A) and SARS-CoV2 specific (B) T cells**. (**A**) Doublets were removed using FSC-A and FSC-A parameters and lymphocytes were gated based on physical parameters (FSC-SSC). Dead cells were excluded using viability stain 780. T cells were identified as CD3+. We then identified CD4+ T cells. Among these, we identified CD154 expressing cells. CD4+CD154+ T cells were then evaluated for IFN-γ, TNF-α and IL-2 expression. (**B**) Doublets were removed using FSC-A and FSC-A parameters and lymphocytes were gated based on physical parameters (FSC-SSC). Dead cells were excluded using viability stain 780. T cells were identified as CD3+. We then identified CD4+ and CD8+ T cells. CD4+ T cells were evaluated for CD154, IFN-γ, TNF-α and IL-2 expression. CD8+ T cells were evaluated for IFN-γ, TNF-α and IL-2 expression.
